# Determinants of protection against SARS-CoV-2 Omicron BA.1 and Delta infections in fully vaccinated outpatients

**DOI:** 10.1101/2023.01.14.23284558

**Authors:** Alvaro Roy, Carla Saade, Laurence Josset, Bénédicte Clément, Florence Morfin, Grégory Destras, Martine Valette, Vinca Icard, Antoine Oblette, Marion Debombourg, Christine Garrigou, Karen Brengel-Pesce, Laurence Generenaz, Kahina Saker, Romain Hernu, Bruno Pozzetto, Bruno Lina, Mary-Anne Trabaud, Sophie Trouillet-Assant, Antonin Bal

## Abstract

**Objectives:** We aimed to evaluate the association between the humoral and cellular immune responses and symptomatic SARS-CoV-2 infection with Delta or Omicron BA.1 variants in fully vaccinated outpatients.

**Methods:** Anti-RBD IgG levels and IFN-γ release were evaluated at PCR-diagnosis of SARS-CoV-2 in 636 samples from negative and positive patients during Delta and Omicron BA.1 periods.

**Results:** Median levels of anti-RBD IgG in positive patients were significantly lower than in negative patients for both variants (*p* < 0.05). The risk of Delta infection was inversely correlated with anti-RBD IgG titres (aOR = 0.63, 95% CI [0.41; 0.95], *p* = 0.03) and it was lower in the hybrid immunity group compared to the homologous vaccination group (aOR = 0.22, 95% CI [0.05; 0.62], *p* = 0.01). In contrast, neither the vaccination scheme nor anti-RBD IgG titers were associated with the risk of BA.1 infection in multivariable analysis. IFN-γ release post-SARS-CoV-2 peptide stimulation was not different between samples from patients infected (either with Delta or Omicron BA.1 variant) or not (*p* = 0.77).

**Conclusions:** Our results show that high circulating levels of anti-RBD IgG and hybrid immunity were independently associated with a lower risk of symptomatic SARS-CoV-2 infection in outpatients with differences according to the infecting variant.

## 1. Introduction

Breakthrough infections are essential to characterise the humoral and cellular immune response, and to search for correlates of protection (CoP) against SARS-CoV-2 infections. The estimation of robust CoP thresholds could help to optimise vaccination strategies but also to accelerate the vaccine authorisation process, and, perhaps, in the future avoid the need of large efficacy trials (1). Both binding and neutralising antibodies have been suggested as major CoP against COVID-19 (1-3). For instance, higher anti-spike IgG, anti-receptor binding domain (RBD) IgG, and neutralising antibody titres have been associated with lower risk of symptomatic disease, although they were not correlated with asymptomatic infections (1); conversely lower levels of neutralising antibodies have been described during the peri-infection period in SARS-CoV-2 breakthrough infections with the B.1.1.7 (Alpha) variant than in matched uninfected controls (4). Similarly, higher anti-spike IgG levels were, more recently, associated with increased protection against Delta variant in BNT162b2 and ChAdOx1-vaccinated individuals (5). The correlation between anti-RBD IgG and neutralising antibodies was high in vaccinated patients against ancestral variants, Alpha, Beta and Delta, however this was not the case for Omicron as it harbours several mutations in the RBD of the spike protein (2, 6). These findings could also imply that the use of anti-RBD IgG as a predictor of protection in Omicron subvariants is compromised, as high levels of anti-RBD IgG may not translate into high titres of neutralising antibodies. This is supported by a cohort study conducted in Denmark in which an association between the level of anti-spike IgG and the risk of breakthrough infection with Delta but not with Omicron BA.1 and BA.2 among fully vaccinated individuals (7). However, other studies have reported an inverse relationship between anti-spike IgG levels and the risk of breakthrough infection with Omicron after the second dose of BNT162b2 vaccine (8), or lower peri-infection anti-spike IgG levels in breakthrough cases compared to matched controls (9). Therefore, further studies in individuals with different vaccination schedules and hybrid immunity are needed to determine whether the CoP for Delta and previous variants of concern (VOCs), such as anti-RBD IgG, are effective with Omicron BA.1 and new Omicron subvariants.

Cellular immunity could play a crucial role in the control of SARS-CoV-2 and its importance may have been relatively underestimated thus far. Kent *et al*. described that current evidence does not support a direct impact of vaccine-induced memory T cells in preventing symptomatic SARS-CoV-2 infection, but their role in preventing severity of infection has not yet been fully explored (10). Unlike humoral immunity, T-cells can provide long-term protection after SARS-CoV-2 infection and after vaccination (11). However, the measurement of cell-based immunity implies greater complexity and costs. Therefore, the evaluation of rapid, high-throughput, and easy to perform cell-based techniques such as an interferon-γ release assay (IGRA) for SARS-CoV-2 is important to determine CoP in order to evaluate current vaccine strategies (12).

The objective of this prospective study was to investigate the association of anti-RBD antibody titre and IGRA positivity with the risk of symptomatic breakthrough infection with Delta or Omicron BA.1 variants in fully vaccinated patients.

## 2. Material and Methods

### 2.1. Study design and population

This prospective test-negative study was conducted at the University Hospital of Lyon, France (*Hospices Civils de Lyon*, HCL) between 23 August 2021 and 2 February 2022. The present study was part of a larger study including healthcare workers (HCWs) and non-HCWs with a full vaccination schedule. Clinical, microbiological, and serological data were collected for all included patients at diagnosis. The inclusion criteria of the larger study were: i) at least 18 years of age, ii) written informed consent for participation, and iii) enrolled in a social security scheme.

For the analysis of the present study, patients were considered if they had i) a full vaccination schedule, defined as at least two doses of COVID-19 vaccines for COVID-19-naïve individuals or as a previous infection followed by at least one vaccine dose; ii) presence of COVID-19 symptoms at the time of inclusion, i.e. at diagnosis; iii) maximum interval of 5 days from symptoms onset to diagnosis; iv) minimum interval of 14 days and maximum interval of 300 days from last immunization (infection or vaccination) and onset of symptoms. Additionally, patients were not considered for the analyses if they had missing information regarding vaccination schemes or regarding previous history of COVID-19.

In our study, hybrid immunity was defined as an immune response induced by an infection and a vaccination, regardless of which immunization occurred first.

### 2.2. Microbiological investigations

Nucleic acid extraction from nasopharyngeal swabs (NPS) was performed on the automated MGISP-960 workstation using MGI Easy Magnetic Kit (MGI Tech, Marupe, Latvia). Quantitative SARS-CoV-2 viral load was determined with qPCR SARS-CoV-2 R-gene kit (bioMérieux, Lyon, France) that includes four quantification standards targeting the SARS-CoV-2 N gene: QS1 to QS4 respectively 2.5.10^6^, 2.5.10^5^, 2.5.10^4^, 2.5.10^3^ copies/mL of a SARS-CoV-2 DNA standard. NSP were also tested using the CELL control R-GENE (bioMérieux) kit to normalise the viral load per 10^4^ cells.

The SARS-CoV-2 variant responsible for infection was confirmed by whole-genome sequencing. The routine SARS-CoV-2 next generation sequencing (NGS) protocol in our laboratory was based on COVIDSeq-TestTM (Illumina, San Diego, CA, USA) using Artic V4 or V4.1 primers as they became available. cDNA synthesis and amplification were performed using the COVID-Seq-TestTM (Illumina). Libraries were prepared with the COVIDSeq-Test (Illumina), and samples were sequenced with 100 bp paired-end reads using the NovaSeq 6000 Sequencing system SP flow cell (13).

### 2.3. Immunological tests

#### 2.3.1. Anti-RBD antibodies

The presence of anti-SARS-CoV-2 antibodies was evaluated in 636 patients. Anti-RBD levels were determined using the Siemens Healthineers (Erlangen, Germany) Atellica^®^ IM SARS-CoV-2 IgG (sCOVG) kit, a fully automated 2-step sandwich immunoassay using indirect chemiluminescent technology, according to the protocol recommended by the manufacturer (14). Antibody levels were reported as binding antibody units (BAU)/ml according to the WHO international standard (15).

#### 2.3.2. Neutralisation assays

We selected a subset of samples (n=35) collected from positive and negative patients during both Delta and Omicron waves. Samples were selected based on high anti-RBD IgG levels (>1000 BAU/mL). A live virus neutralisation test was performed against 19A, Delta, and Omicron BA.1 isolates using these samples. Briefly, each serum specimen was diluted 1:10 in culture medium (Dulbecco’s Modified Eagle medium), then heated for 30 min at 56°C to avoid activation of complement. Serial 2-fold dilutions of the serum were mixed at equal volume with the live SARS-CoV-2 isolate, previously diluted to the desired concentration. A plate was prepared in parallel to determine the viral titre. After gentle shaking and contact for 30 min at room temperature, 150μL of the mix was transferred into a 96-well microplate covered with Vero E6 cells. The plates were incubated at 37°C in a 5% CO_2_ atmosphere. Measurements were obtained microscopically 4-5 days later when the cytopathic effect of the virus control reached 100 Tissue Culture Infectious Dose 50 (TCID_50_)/150μL. The serum was considered to have a protective effect if more than 50% of the cells were preserved. The neutralisation titre was expressed as the inverse of the highest serum dilution that allowed protection of the cells. Global Initiative on Sharing Avian Influenza Data (GISAID) accession numbers are EPI_ISL_1707038, EPI_ISL_1904989 and EPI_ISL_7608613 for the 19A, Delta and Omicron BA.1 isolates used respectively. These experiments were performed in a biosafety level 3 laboratory as previously described (16, 17). Fold changes were calculated for positive neutralisation titres against both Delta and Omicron BA.1 isolates by comparing titres obtained for these isolates with those obtained against the 19A isolate.

### 2.4. Interferon-gamma release assay (IGRA)

The cellular response was investigated using an interferon-gamma (IFN-γ) release assay (IGRA) as previously described (18) using the VIDAS^®^ COVID stimulation and VIDAS^®^ 9IFN Research Use Only (RUO) kits (bioMérieux). These experiments were performed on a subset of samples from 243 included patients. In brief, whole blood was stimulated with a restricted pool of peptides (RPP) specific to SARS-CoV-2 structural proteins. The supernatant was then collected 22 hours post-stimulation and IFN-γ release was quantified using an automated VIDAS^®^ ELISA test to determine IFN-γ levels for each sample. According to manufacturer’s instructions, samples with IFN-γ release ≥ 0.13 IU/mL were considered IGRA-positive, if not then samples were considered IGRA-negative.

### 2.5. Statistical analysis

Comparisons of quantitative results between two groups were conducted using the Mann-Whitney U test (after checking the absence of normality of the data using the Kolmogorov– Smirnov test) and the associations of categorical variables were analysed using the Pearson’s Chi-squared or Fisher’s exact tests when appropriated. The correlation between viral load and anti-RBD IgG levels was analysed using the Spearman correlation test.

The variables that were significantly associated in the univariate analysis with SARS-CoV-2 infection during Delta period, in order to check the influence of confounding factors and interactions, were then controlled with multivariable logistic regression to obtain adjusted odds ratios (aOR) with 95% confidence intervals [95% CI]. After constructing the maximum multivariable logistic regression model for Delta infection, a confounder remained in the final estimated model if the coefficient for anti-RBD IgG levels and hybrid vaccine regimen changed by more than ten percent when the potential confounder was removed. Analyses were conducted using GraphPad Prism^®^ software (version 8; GraphPad software, La Jolla, CA, USA) and R software, version 4.1.3 (R Foundation for Statistical Computing, Vienna, Austria).

## 3. Results

### 3.1. Patient characteristics and symptoms

Between 23 August 2021 and 2 February 2022, a total of 852 patients were enrolled. Among them, 186 patients were not considered for the analyses presented herein. Patients were divided into two groups according to the predominance of the Delta or Omicron variants determined by SARS-CoV-2 whole genome sequencing results obtained from samples of patients enrolled in the present study (Suppl. Fig. 1). The Delta period was set between 23 August 2021 (week 34) and 12 December 2021 (week 49), while the Omicron period was set from 27 December 2021 (week 52) to 2 February 2022 (week 5). All patients recruited during the weeks 50 and 51 were excluded from the analysis due to co-circulation of both variants (Fig.1). Finally, 380 patients were included in the Delta period and 256 patients in the Omicron period (Fig. 1).

**Fig. 1.**
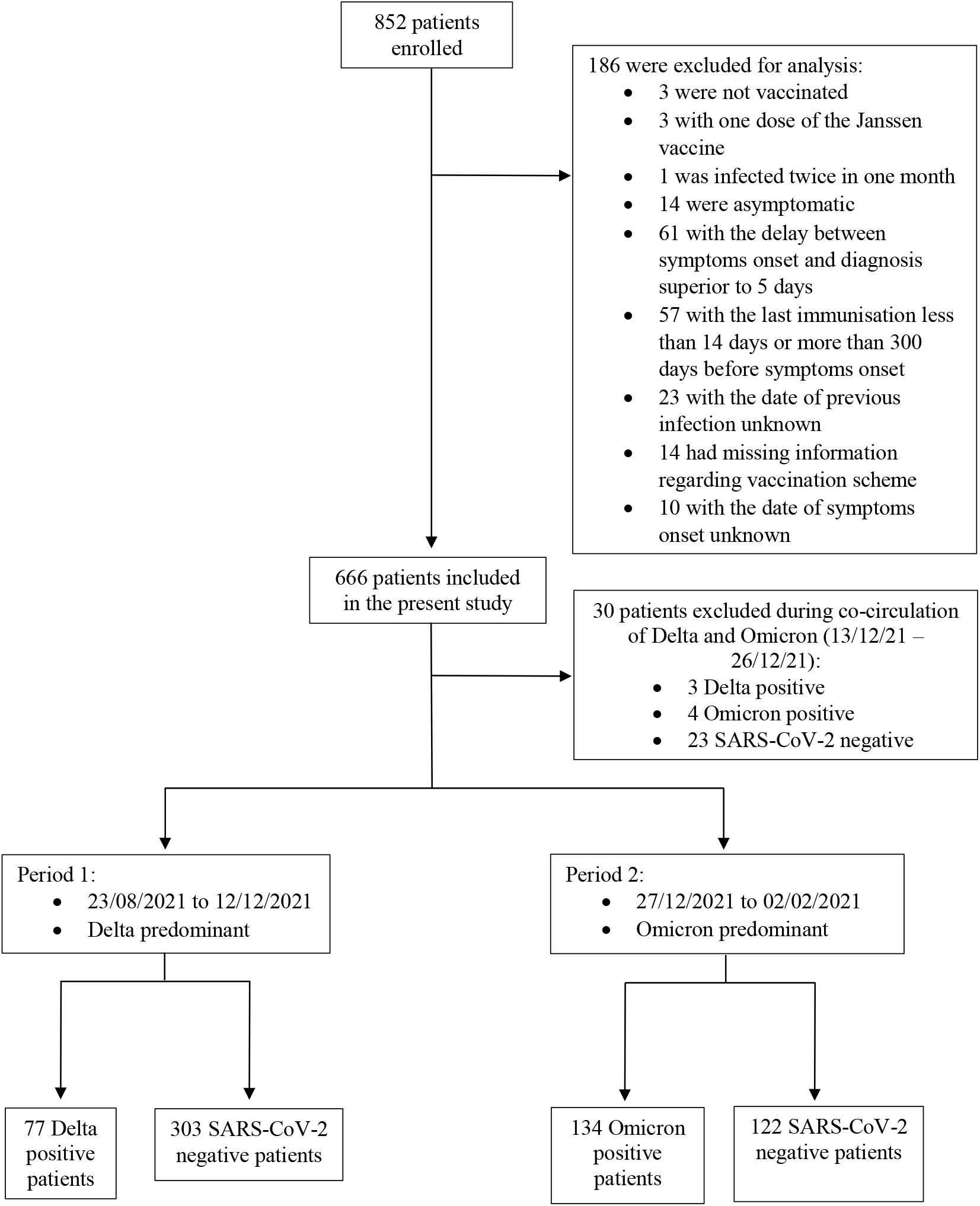
Study flow chart.

We did not observe significant differences in the demographic characteristics between SARS-CoV-2 positive and negative patients in both Delta and Omicron periods, nor between positive patients between periods, except for higher median BMI in Omicron SARS-CoV-2 negative patients compared to Omicron positives (Table 1).

**Table 1.**
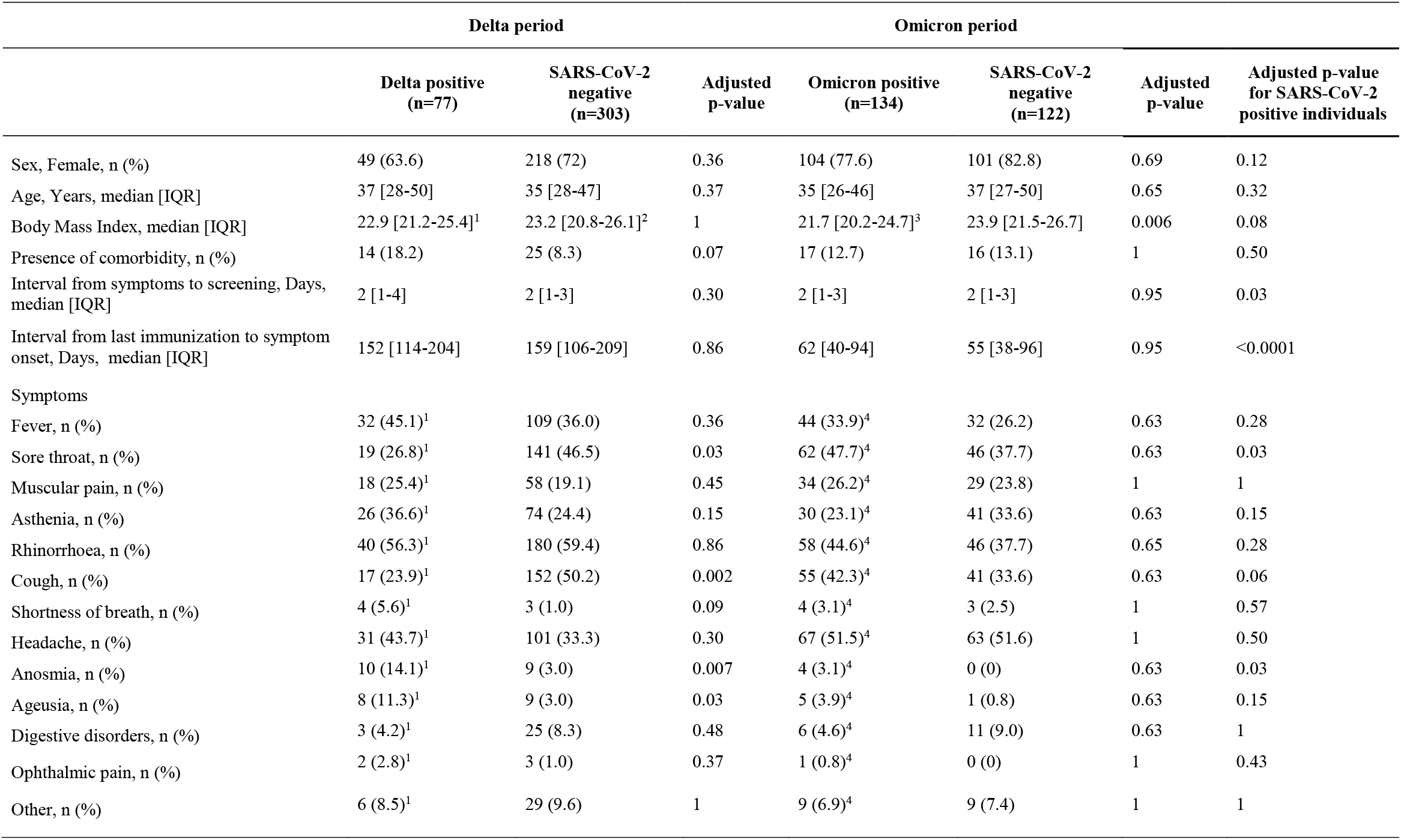
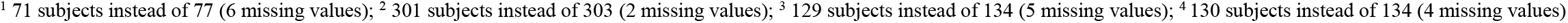
Demographic and clinical characteristics of patients from Delta and Omicron periods.

|  | Delta period |  |  | Omicron period |  |  |  |
| --- | --- | --- | --- | --- | --- | --- | --- |
|  | Delta positive<br>(n=77) | SARS-CoV-2<br>negative<br>(n=303) | Adjusted<br>p-value | Omicron positive<br>(n=134) | SARS-CoV-2<br>negative<br>(n=122) | Adjusted<br>p-value | Adjusted p-value<br>for SARS-CoV-2<br>positive individuals |
| Sex, Female, n (%) | 49 (63.6) | 218 (72) | 0.36 | 104 (77.6) | 101 (82.8) | 0.69 | 0.12 |
| Age, Years, median [IQR] | 37 [28-50] | 35 [28-47] | 0.37 | 35 [26-46] | 37 [27-50] | 0.65 | 0.32 |
| Body Mass Index, median [IQR] | 22.9 [21.2-25.4]<sup>1</sup> | 23.2 [20.8-26.1]<sup>2</sup> | 1 | 21.7 [20.2-24.7]<sup>3</sup> | 23.9 [21.5-26.7] | 0.006 | 0.08 |
| Presence of comorbidity, n (%) | 14 (18.2) | 25 (8.3) | 0.07 | 17 (12.7) | 16 (13.1) | 1 | 0.50 |
| Interval from symptoms to screening, Days, median [IQR] | 2 [1-4] | 2 [1-3] | 0.30 | 2 [1-3] | 2 [1-3] | 0.95 | 0.03 |
| Interval from last immunization to symptom onset, Days, median [IQR] | 152 [114-204] | 159 [106-209] | 0.86 | 62 [40-94] | 55 [38-96] | 0.95 | <0.0001 |
| Symptoms |  |  |  |  |  |  |  |
| Fever, n (%) | 32 (45.1)<sup>1</sup> | 109 (36.0) | 0.36 | 44 (33.9)<sup>4</sup> | 32 (26.2) | 0.63 | 0.28 |
| Sore throat, n (%) | 19 (26.8)<sup>1</sup> | 141 (46.5) | 0.03 | 62 (47.7)<sup>4</sup> | 46 (37.7) | 0.63 | 0.03 |
| Muscular pain, n (%) | 18 (25.4)<sup>1</sup> | 58 (19.1) | 0.45 | 34 (26.2)<sup>4</sup> | 29 (23.8) | 1 | 1 |
| Asthenia, n (%) | 26 (36.6)<sup>1</sup> | 74 (24.4) | 0.15 | 30 (23.1)<sup>4</sup> | 41 (33.6) | 0.63 | 0.15 |
| Rhinorrhoea, n (%) | 40 (56.3)<sup>1</sup> | 180 (59.4) | 0.86 | 58 (44.6)<sup>4</sup> | 46 (37.7) | 0.65 | 0.28 |
| Cough, n (%) | 17 (23.9)<sup>1</sup> | 152 (50.2) | 0.002 | 55 (42.3)<sup>4</sup> | 41 (33.6) | 0.63 | 0.06 |
| Shortness of breath, n (%) | 4 (5.6)<sup>1</sup> | 3 (1.0) | 0.09 | 4 (3.1)<sup>4</sup> | 3 (2.5) | 1 | 0.57 |
| Headache, n (%) | 31 (43.7)<sup>1</sup> | 101 (33.3) | 0.30 | 67 (51.5)<sup>4</sup> | 63 (51.6) | 1 | 0.50 |
| Anosmia, n (%) | 10 (14.1)<sup>1</sup> | 9 (3.0) | 0.007 | 4 (3.1)<sup>4</sup> | 0 (0) | 0.63 | 0.03 |
| Ageusia, n (%) | 8 (11.3)<sup>1</sup> | 9 (3.0) | 0.03 | 5 (3.9)<sup>4</sup> | 1 (0.8) | 0.63 | 0.15 |
| Digestive disorders, n (%) | 3 (4.2)<sup>1</sup> | 25 (8.3) | 0.48 | 6 (4.6)<sup>4</sup> | 11 (9.0) | 0.63 | 1 |
| Ophthalmic pain, n (%) | 2 (2.8)<sup>1</sup> | 3 (1.0) | 0.37 | 1 (0.8)<sup>4</sup> | 0 (0) | 1 | 0.43 |
| Other, n (%) | 6 (8.5)<sup>1</sup> | 29 (9.6) | 1 | 9 (6.9)<sup>4</sup> | 9 (7.4) | 1 | 1 |

<sup>1</sup> 71 subjects instead of 77 (6 missing values); <sup>2</sup> 301 subjects instead of 303 (2 missing values); <sup>3</sup> 129 subjects instead of 134 (5 missing values); <sup>4</sup> 130 subjects instead of 134 (4 missing values)

The main symptoms reported by SARS-CoV-2 positive patients during the Delta period were rhinorrhoea (56.3%), fever (45.1%), and headache (43.7%). Anosmia and ageusia were significantly more frequently reported by positive patients while sore throat and cough were twice less frequently reported by positive than negative patients (*p* < 0.05). Regarding the Omicron period, the main symptoms reported by SARS-CoV-2 positive patients were headache (51.5%), sore throat (47.7%), rhinorrhoea (44.6%) and cough (42.3%); there was no significant difference in the frequency of any symptom between positive and negative patients. Sore throat was significantly more frequently reported among Omicron-positive patients compared to Delta-positive patients (respectively, 47.7% vs 26.8%, *p* = 0.03). Conversely, anosmia was more frequent in Delta-positive patients (14.1%) than in Omicron-positive patients (3.1%, *p* = 0.03).

There was no significant difference in the interval from last immunization (previous infection or vaccination) to the onset of symptoms between SARS-CoV-2 positive and negative patients in each period; Omicron-positive patients had a shorter interval since last immunization to the onset of symptoms compared to Delta-positive patients (median 62 days vs median 152 days, *p* < 0.0001; Table 1). Overall, a greater proportion of patients had three rather than two immunizations during the Omicron period (80.5%) than in the Delta period (10.8%, *p* < 0.001; Table 2).

**Table 2.**
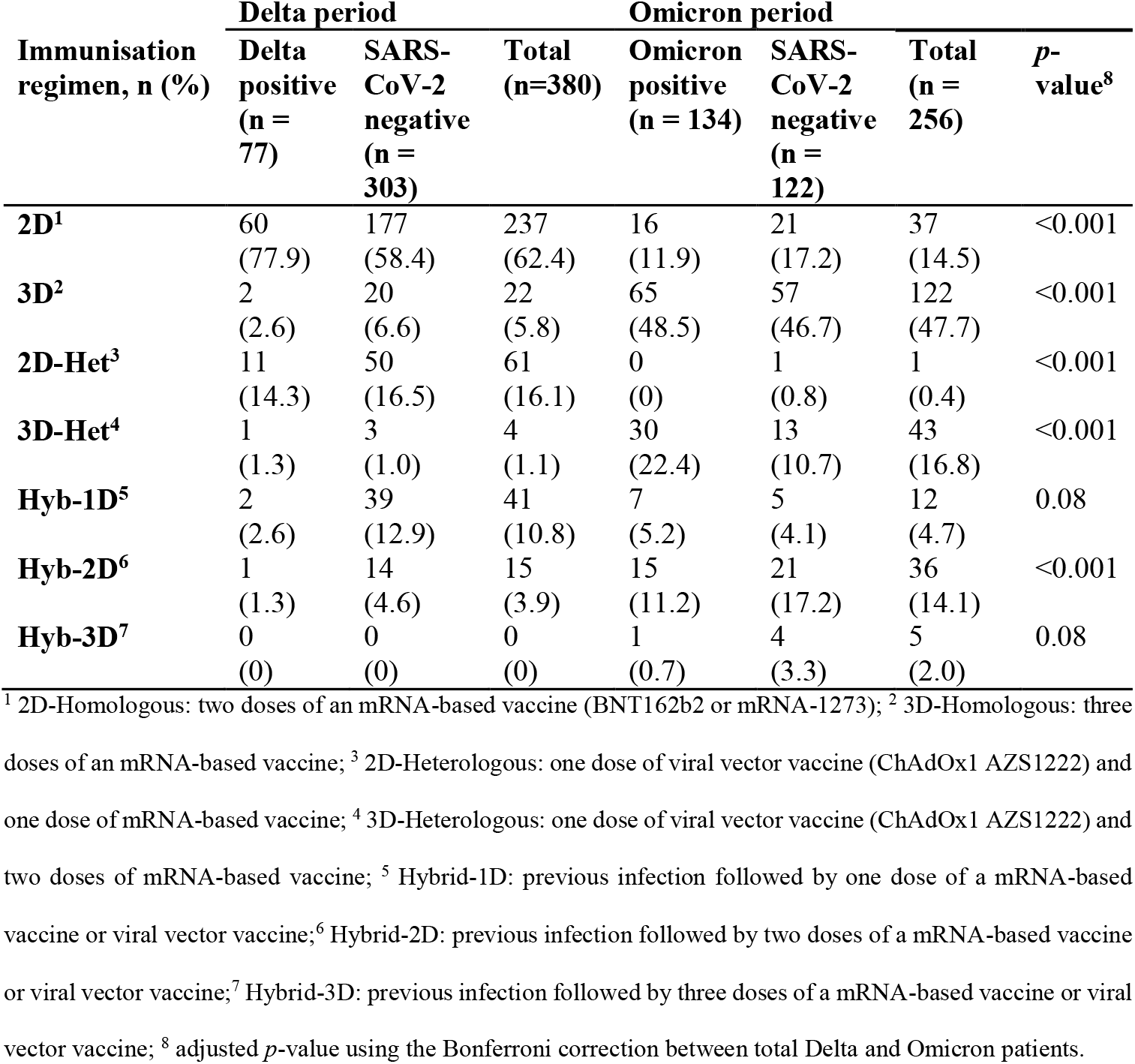
Number and percentage of SARS-CoV-2 positive and negative patients according to vaccination regimen stratified by period.

| Immunisation regimen, n (%) | Delta period |  |  | Omicron period |  |  | *p*-value<sup>8</sup> |
| --- | --- | --- | --- | --- | --- | --- | --- |
|  | Delta positive (n = 77) | SARS-CoV-2 negative (n = 303) | Total (n=380) | Omicron positive (n = 134) | SARS-CoV-2 negative (n = 122) | Total (n = 256) |  |
| **2D<sup>1</sup>** | 60<br>(77.9) | 177<br>(58.4) | 237<br>(62.4) | 16<br>(11.9) | 21<br>(17.2) | 37<br>(14.5) | <0.001 |
| **3D<sup>2</sup>** | 2<br>(2.6) | 20<br>(6.6) | 22<br>(5.8) | 65<br>(48.5) | 57<br>(46.7) | 122<br>(47.7) | <0.001 |
| **2D-Het<sup>3</sup>** | 11<br>(14.3) | 50<br>(16.5) | 61<br>(16.1) | 0<br>(0) | 1<br>(0.8) | 1<br>(0.4) | <0.001 |
| **3D-Het<sup>4</sup>** | 1<br>(1.3) | 3<br>(1.0) | 4<br>(1.1) | 30<br>(22.4) | 13<br>(10.7) | 43<br>(16.8) | <0.001 |
| **Hyb-1D<sup>5</sup>** | 2<br>(2.6) | 39<br>(12.9) | 41<br>(10.8) | 7<br>(5.2) | 5<br>(4.1) | 12<br>(4.7) | 0.08 |
| **Hyb-2D<sup>6</sup>** | 1<br>(1.3) | 14<br>(4.6) | 15<br>(3.9) | 15<br>(11.2) | 21<br>(17.2) | 36<br>(14.1) | <0.001 |
| **Hyb-3D<sup>7</sup>** | 0<br>(0) | 0<br>(0) | 0<br>(0) | 1<br>(0.7) | 4<br>(3.3) | 5<br>(2.0) | 0.08 |

<sup>1</sup> 2D-Homologous: two doses of an mRNA-based vaccine (BNT162b2 or mRNA-1273); <sup>2</sup> 3D-Homologous: three doses of an mRNA-based vaccine; <sup>3</sup> 2D-Heterologous: one dose of viral vector vaccine (ChAdOx1 AZS1222) and one dose of mRNA-based vaccine; <sup>4</sup> 3D-Heterologous: one dose of viral vector vaccine (ChAdOx1 AZS1222) and two doses of mRNA-based vaccine; <sup>5</sup> Hybrid-1D: previous infection followed by one dose of a mRNA-based vaccine or viral vector vaccine; <sup>6</sup> Hybrid-2D: previous infection followed by two doses of a mRNA-based vaccine or viral vector vaccine; <sup>7</sup> Hybrid-3D: previous infection followed by three doses of a mRNA-based vaccine or viral vector vaccine; <sup>8</sup> adjusted *p*-value using the Bonferroni correction between total Delta and Omicron patients.

### 3.2. Association of anti-RBD IgG levels and risk of infection

As Delta and Omicron waves occurred at different times during the vaccination campaign, the median anti-RBD IgG levels at diagnosis differed between periods. The median anti-RBD IgG levels were significantly higher in positive patient samples collected during the Omicron period compared to samples collected during the Delta period (*p* < 0.0001, Fig. 2), with titres approximately 7.7-fold higher. During the Delta period, the median [IQR] level of anti-RBD IgG among positive patients was 235 [108-613] BAU/mL, which was significantly lower than that of negative patients (374 [163-1048] BAU/mL, *p* = 0.0045; Fig. 2). Likewise, during the Omicron period, the median [IQR] anti-RBD IgG level was 1881 [941-2533] BAU/mL in positive patients and 2295 [1220-3374] BAU/mL in negative patients, although the difference was less marked during this period (*p* = 0.02; Fig. 2).

**Figure 2.**
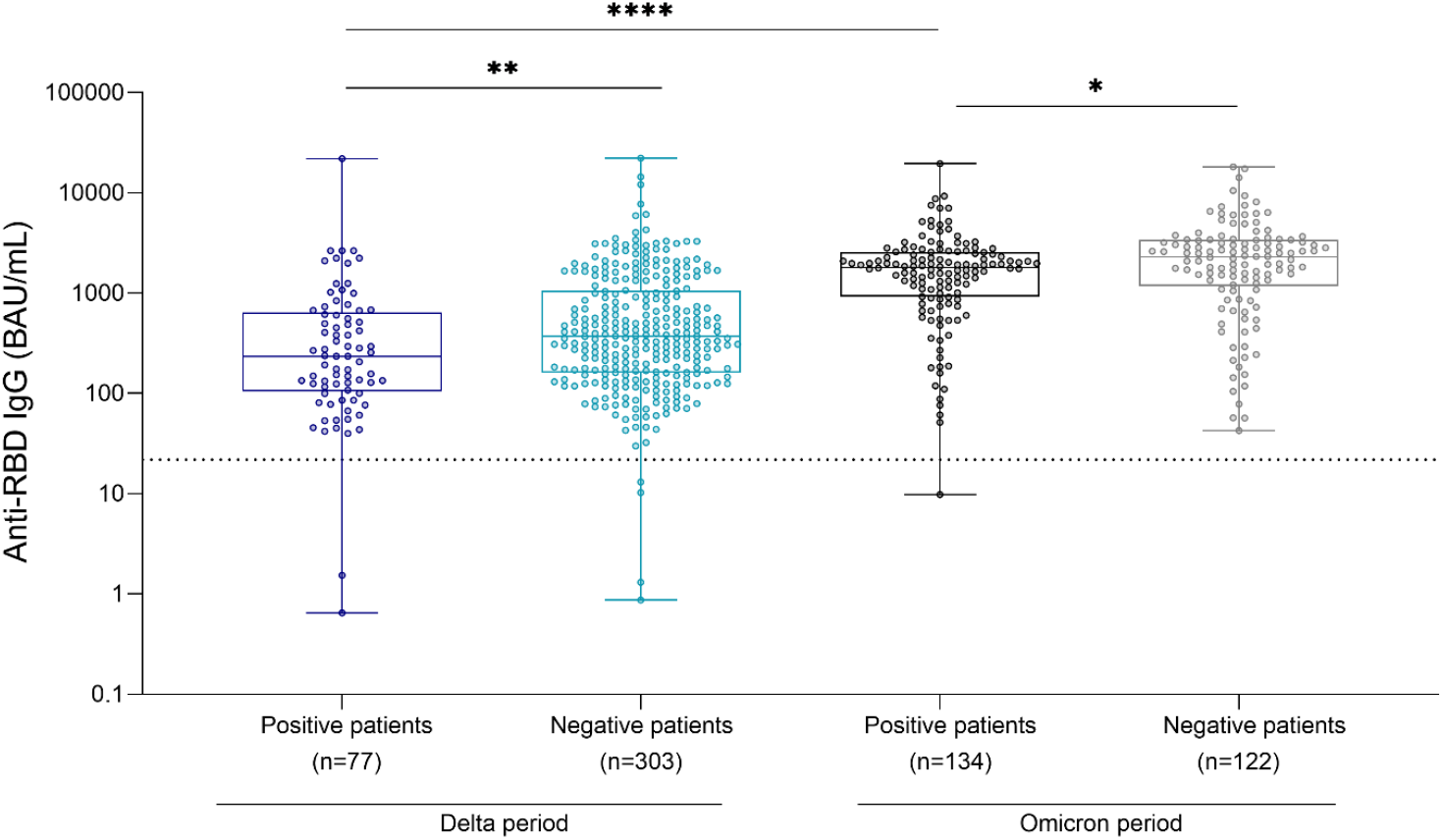
Comparison of anti-RBD IgG levels between SARS-CoV-2 positive and negative patients during Delta and Omicron periods. A chemiluminescence immunoassay (CLIA) was performed in order to determine anti-RBD IgG levels, expressed in binding antibody units (BAU)/mL (n=636). The dotted line represents the threshold of positivity (≥ 21.8 BAU/mL). Box plots represent the median and interquartile range [IQR], and whiskers represent the range. Comparisons were made using the Mann-Whitney test. \**p*=0.05; \*\**p*<0.01; \*\*\*\**p*<0.0001.

The odds of infection during Delta period decrease by 0.45 for each 10-fold increase in anti-RBD antibody level (OR = 0.55, 95% CI [0.36; 0.83], *p* = 0.005) and by 0.82 in patients with hybrid regimen compared to homologous vaccination (OR = 0.18, 95% CI [0.04; 0.51], *p* = 0.005). No association of Delta infection with sex, age and time since last immunization was observed. In case of Omicron BA.1 infection, only heterologous vaccination compared to homologous vaccination was slightly associated with higher risk of infection in the univariate analysis (OR = 2.06, 95% CI [1.03-4.29]; Fig. 3A), but none of the variables were significantly associated with the risk of Omicron infection in multivariate analysis (Fig. 3B). Conversely, in the multivariable logistic regression analysis, we found that a log10 increase in anti-RBD IgG level (aOR = 0.63, 95% CI [0.41; 0.95], *p* = 0.03) and the hybrid regimen (aOR = 0.22, 95% CI [0.05; 0.62], *p* = 0.01) significantly decreased the risk of Delta infection (Table 3).

**Table 3.**
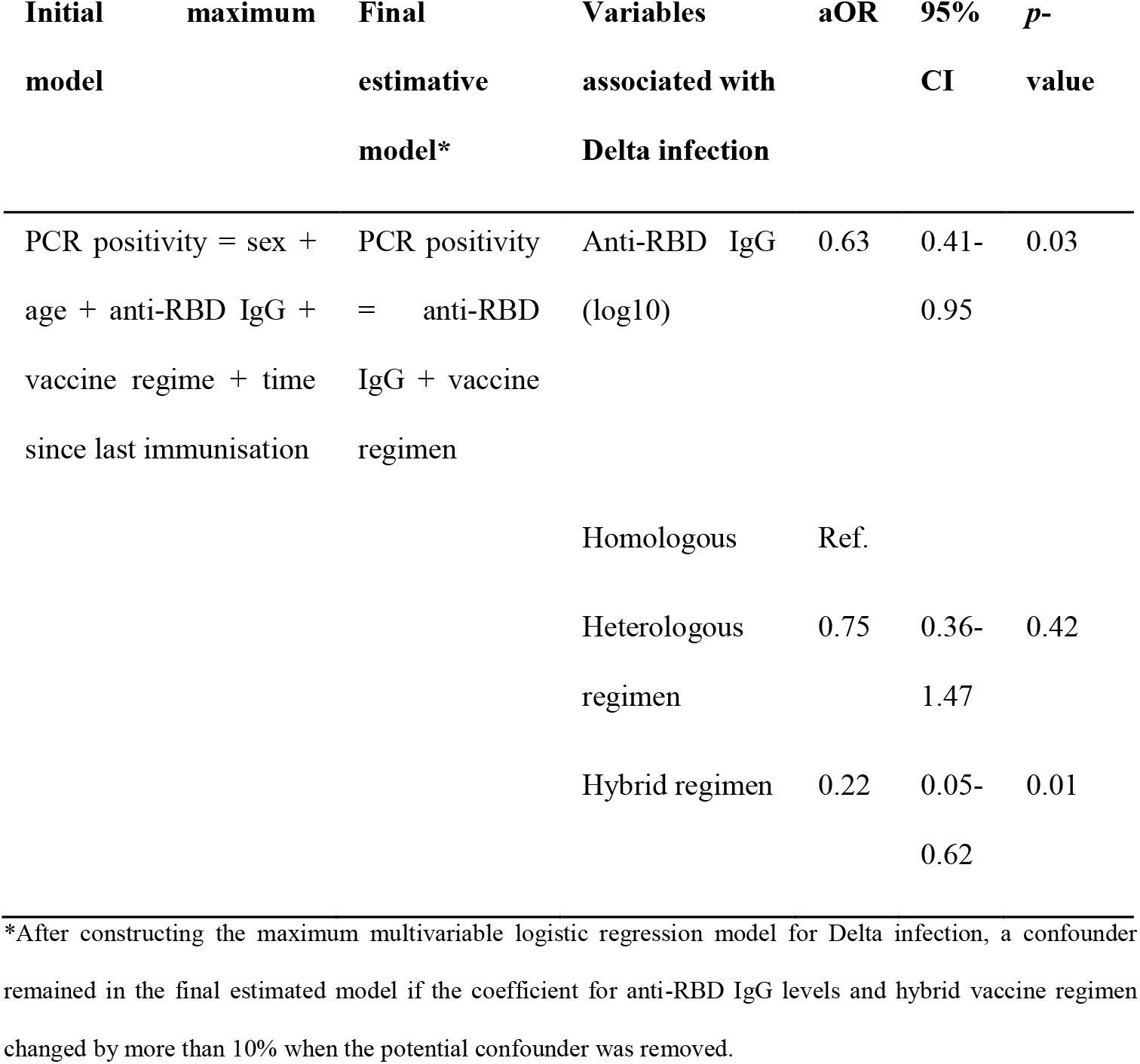
Multivariable analysis of the risk of SARS-CoV-2 infection during the Delta period.

**Figure 3.**
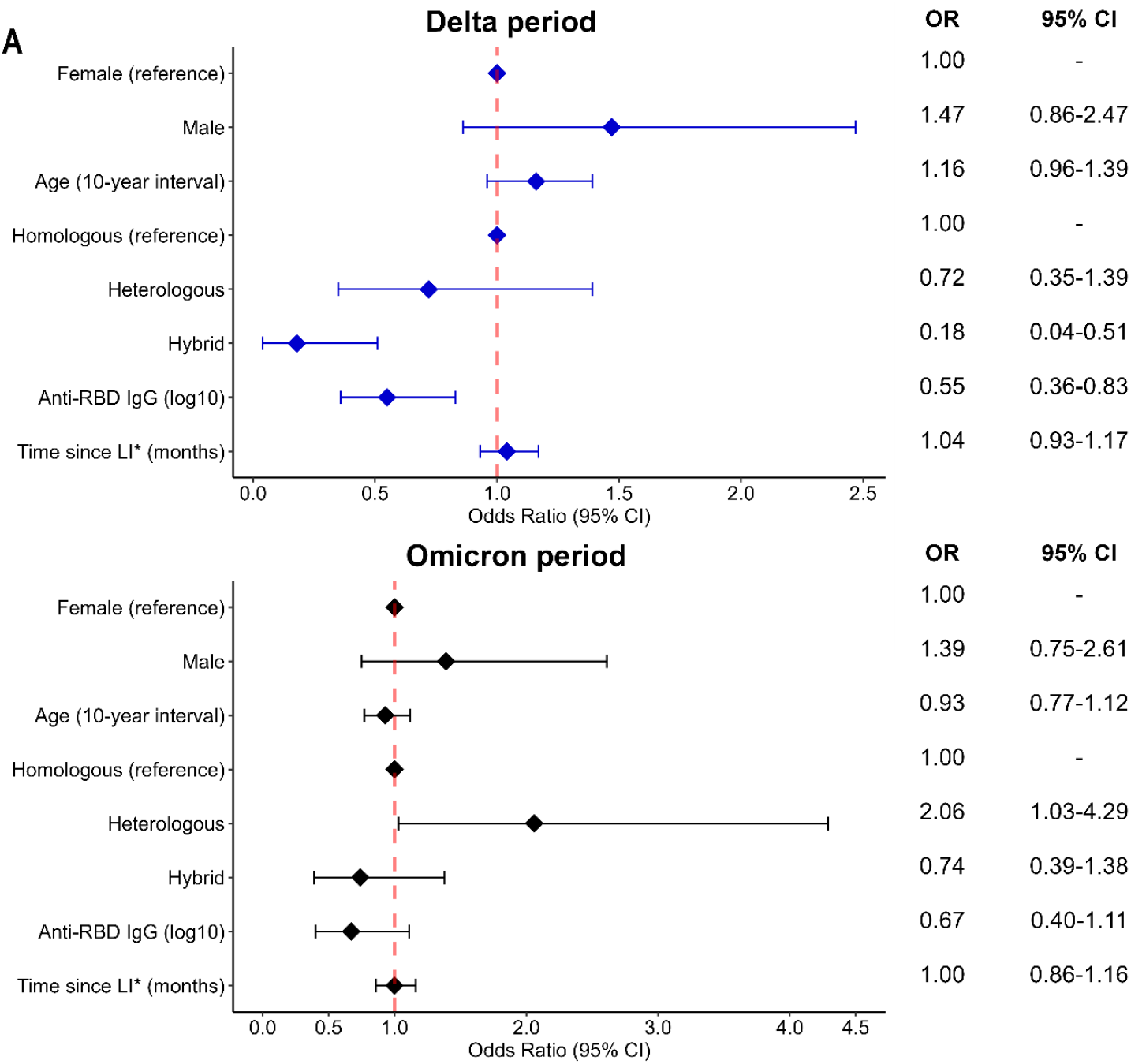
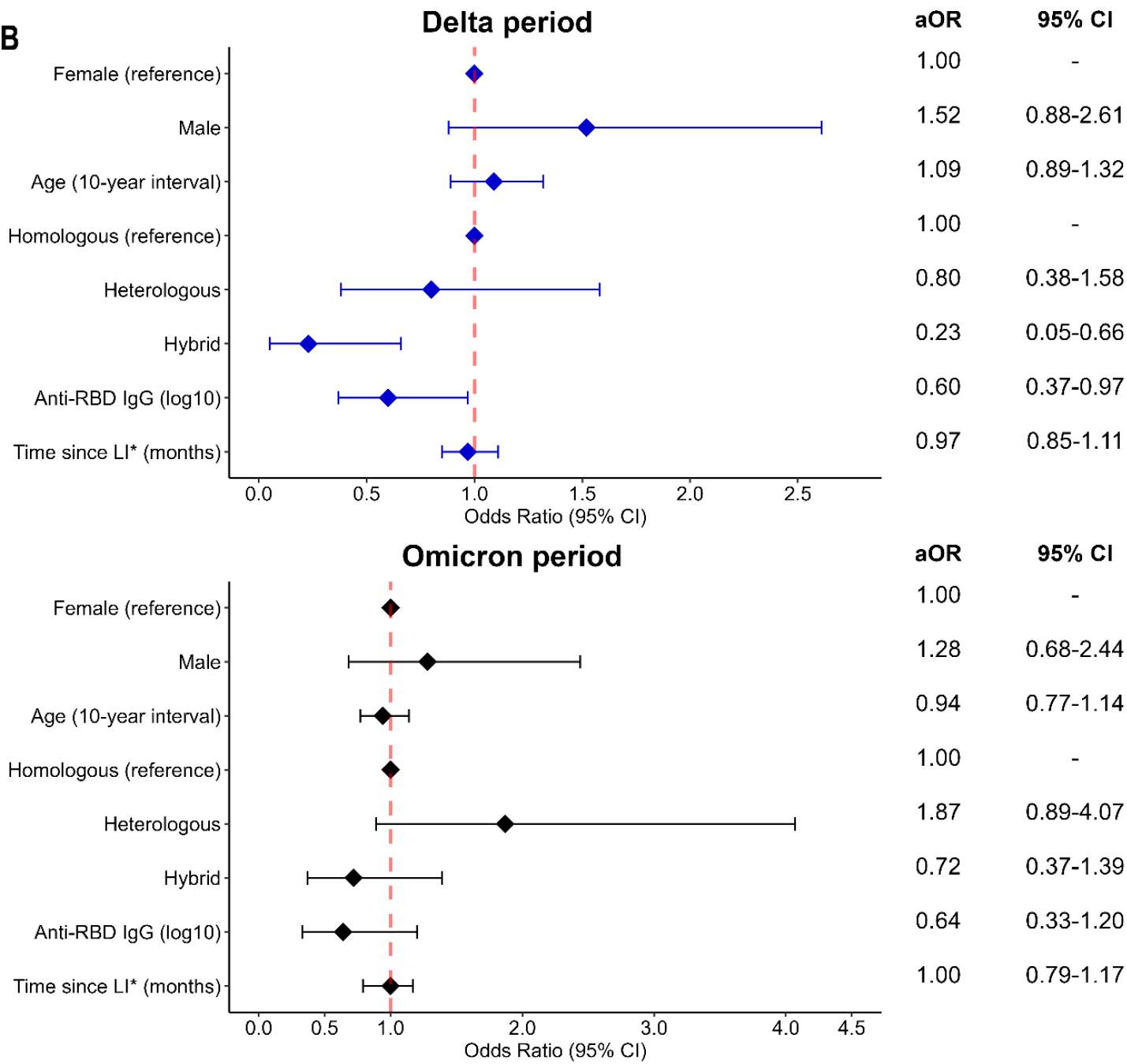
Forest plots showing the crude (A) and adjusted (B) odds ratios for SARS-CoV-2 infection according to sex, age, immunisation status, anti-RDB IgG levels and time since last immunisation in the Delta and Omicron periods. **\***Months since last immunisation

When we studied the association of anti-RBD IgG levels and the risk of Delta and Omicron infection stratified by vaccination regimen we observed the lowest risk of infection as anti-RBD IgG levels increased in the hybrid group compared to heterologous and homologous groups in the case of Delta (Fig. 4). In the case of Omicron BA.1 infection a significant decrease in the risk of infection was observed only for the hybrid group (OR = 0.08, 95% CI [0.01; 0.37], *p* = 0.004; Suppl. Table 1), mainly between 1000 and 10000 BAU/ml (Fig. 4F).

**Figure 4.**
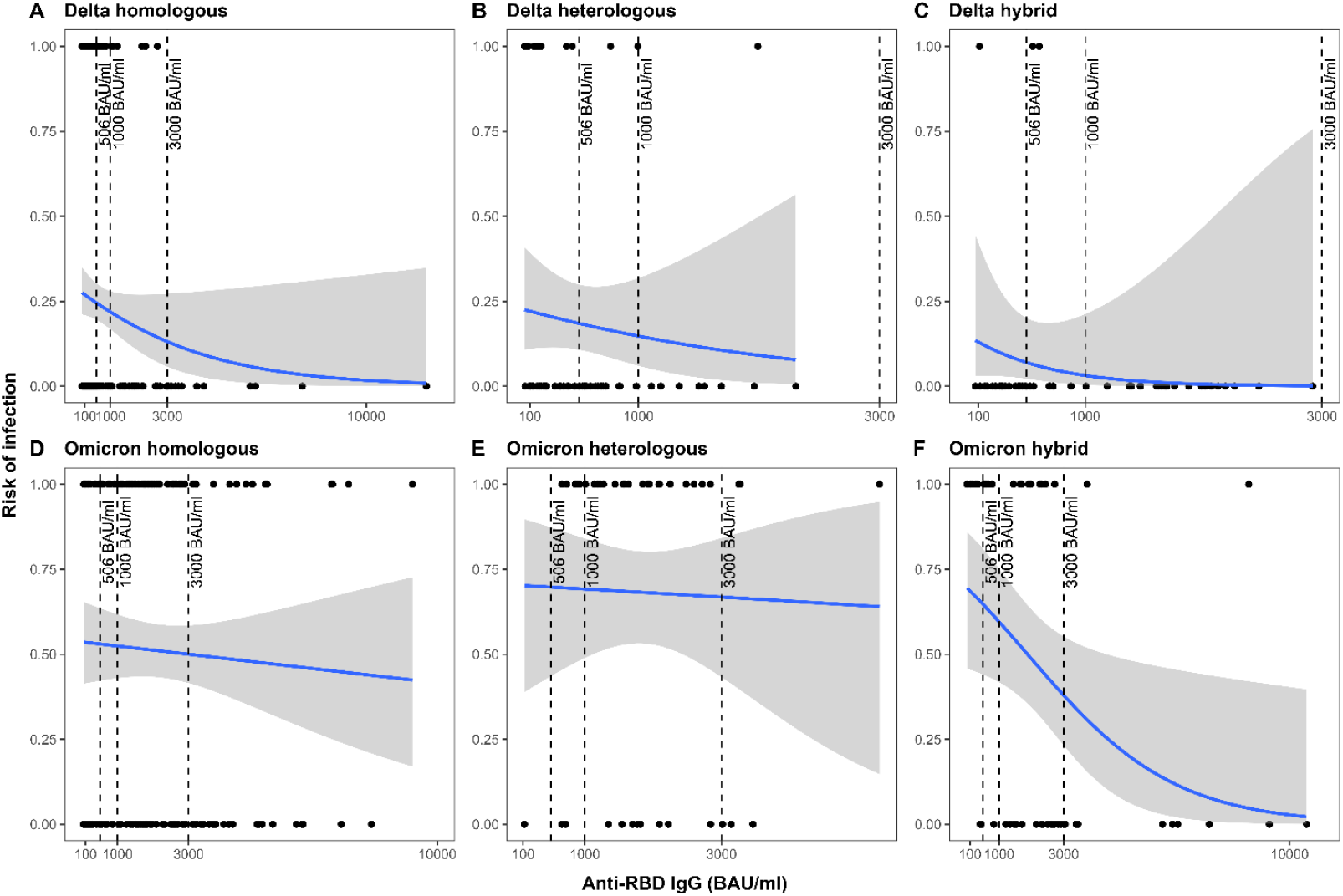
Risk of infection during Delta and Omicron periods according to anti-RBD IgG levels stratified by immunization regimen. Blue solid lines indicate the probability of infection and grey shaded areas the 95% confidence interval. *The threshold of 506 BAU/ml was estimated by Feng *et al*. (1) for a vaccine efficacy of 80% against symptomatic Alpha (B.1.1.7) infections.

When applying the threshold of 506 BAU/ml reported by Feng *et al*. (2) the risk of Delta infection was 24% (95% CI [19.1; 29.7]) in the homologous group, 18.5% (95% CI [10.7; 29.8]) in the heterologous group, and 7% (95% CI [2.2; 20]) in the hybrid group. Given that patients in the Omicron period had higher initial anti-RBD IgG titres due to the recent third dose, when applying the cut-off point reported by Feng *et al*. (2), the predicted risk of infection in the hybrid group was 64.8% (95% CI [44.2; 81.0]), not dropping below 20% until the titre was above 5000 BAU/ml (Fig. 4F).

### 3.3. Neutralisation assays

We selected a subset of sera (n = 35) from SARS-CoV-2 negative and positive patients from both periods presenting high levels of anti-RBD IgG to explore whether despite similar levels of anti-RBD IgG there was a difference in SARS-CoV-2 neutralisation capacity between negative and positive patients. Among the tested sera, 10 were from positive patients and 9 from negative patients from the Delta period, and 7 were from positive patients and 9 from negative patients during the Omicron period. Among sera from the Delta period, the median [IQR] anti-RBD IgG levels was 2166 [1247-2652] BAU/mL for positive patient samples and 2102 [1471-2824] BAU/mL for negative patient samples (Fig. 5A); among those from the Omicron period these were 7054 [5161-8732] BAU/mL for positive patient samples and 6034 [5025-7025] BAU/mL for negative patient samples (Fig. 5B). The median neutralising antibody titres against the corresponding period isolate were not significantly different between positive and negative patients (*p* > 0.05, Fig. 5).

**Figure 5.**
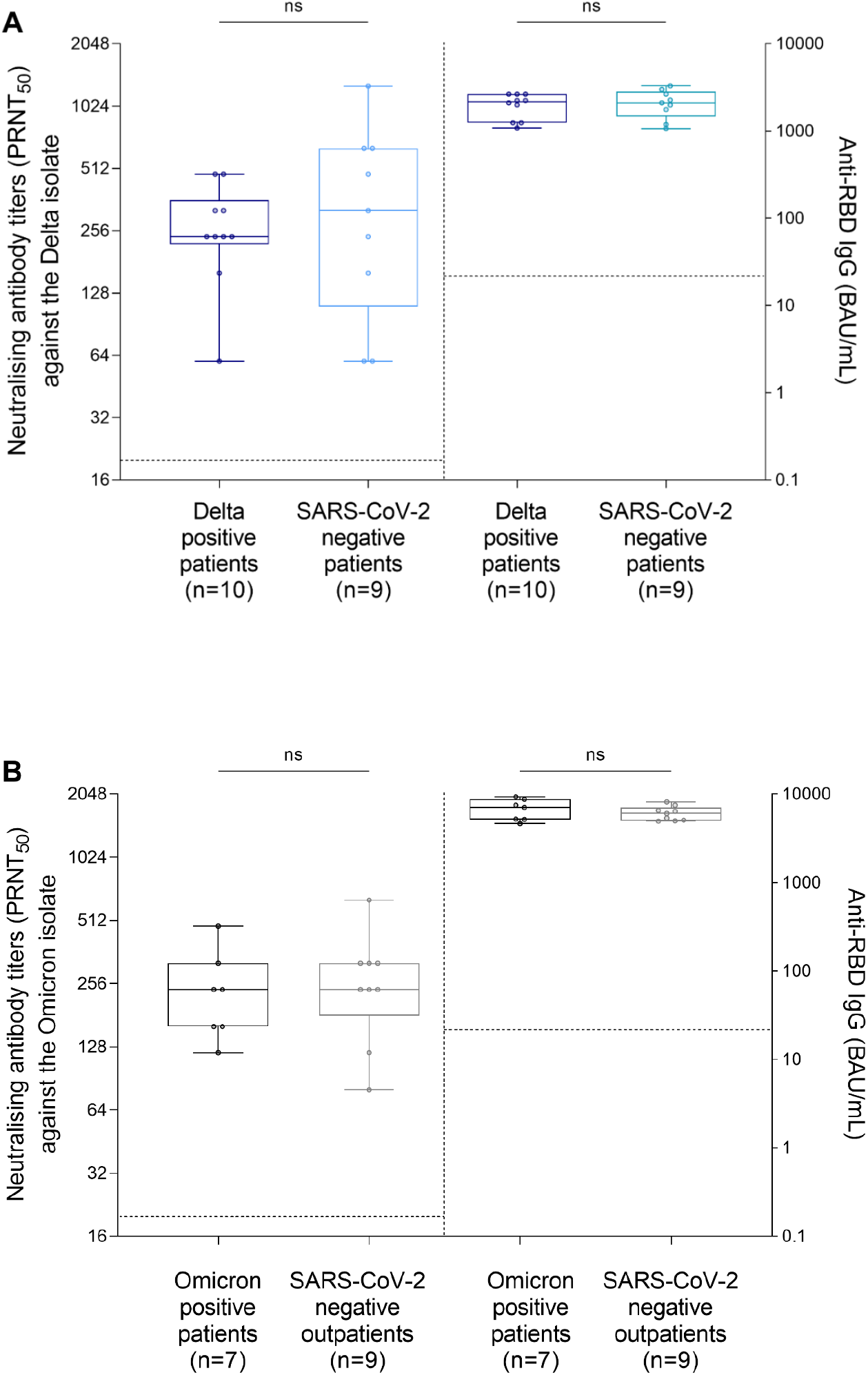
Comparison of anti-RBD IgG levels and neutralising antibody titres between SARS-CoV-2 positive and negative patients samples from both periods. A 50% plaque reduction neutralisation test (PRNT_50_) was conducted to determine the neutralisation titre of 35 sera with high anti-RBD IgG levels corresponding to patients from both periods, against the live Delta and Omicron BA.1 isolates. Anti-RBD IgG levels and neutralisation titres from Delta patients against the Delta isolate (A) and anti-RBD IgG levels and neutralisation titres from Omicron patients against Omicron BA.1 isolate (B). Dotted lines represent the threshold of positivity (≥20 for neutralising titres and ≥21.8 BAU/mL for anti-RBD IgG levels). Comparisons were made using the Mann-Whitney test, ns: non-significant.

Among the tested sera, 100% had neutralizing activity against the 19A isolates, 100% against Delta isolates, and 80% against Omicron BA.1 isolates. The median [IQR] fold reduction in neutralisation titres against the Delta isolate compared to the 19A isolate was two [1.3-2.6] for the Delta and two [1.1-2] for the Omicron period samples, positive and negative patients combined; for the Omicron BA.1 isolate there was a median twelve-fold reduction (IQR [6-24]) compared to the 19A for the Delta period samples and a median eight-fold reduction (IQR [5.3-12]) for Omicron period samples (Suppl. Fig. 2).

### 3.4. Interferon-gamma release assay (IGRA)

In total 243 samples were used for the IGRA; 24 from positive and 76 from negative patients from the Delta period, and 78 from positive and 65 from negative patients from the Omicron period. Stratified according to SARS-CoV-2 positivity status, at least 62.5% of samples in both periods were IGRA-positive (Fig. 6), and there was no significant difference in the proportion of IGRA-positive samples (*p =* 0.77). There was a trend towards a higher median anti-RBD IgG level among IGRA-positive samples compared to IGRA-negative samples among positive patients for both periods as well as among negative patients in the Delta period (at least 1.4 fold higher), and this difference was significant for negative patients in the Omicron period (*p* < 0.01; Suppl. Fig. 3). Moreover, the proportion of IGRA positive samples was higher in the patients with hybrid immunity compared to those with immunizations based on vaccination alone (*p* = 0.02, Suppl. Table 2).

**Figure 6.**
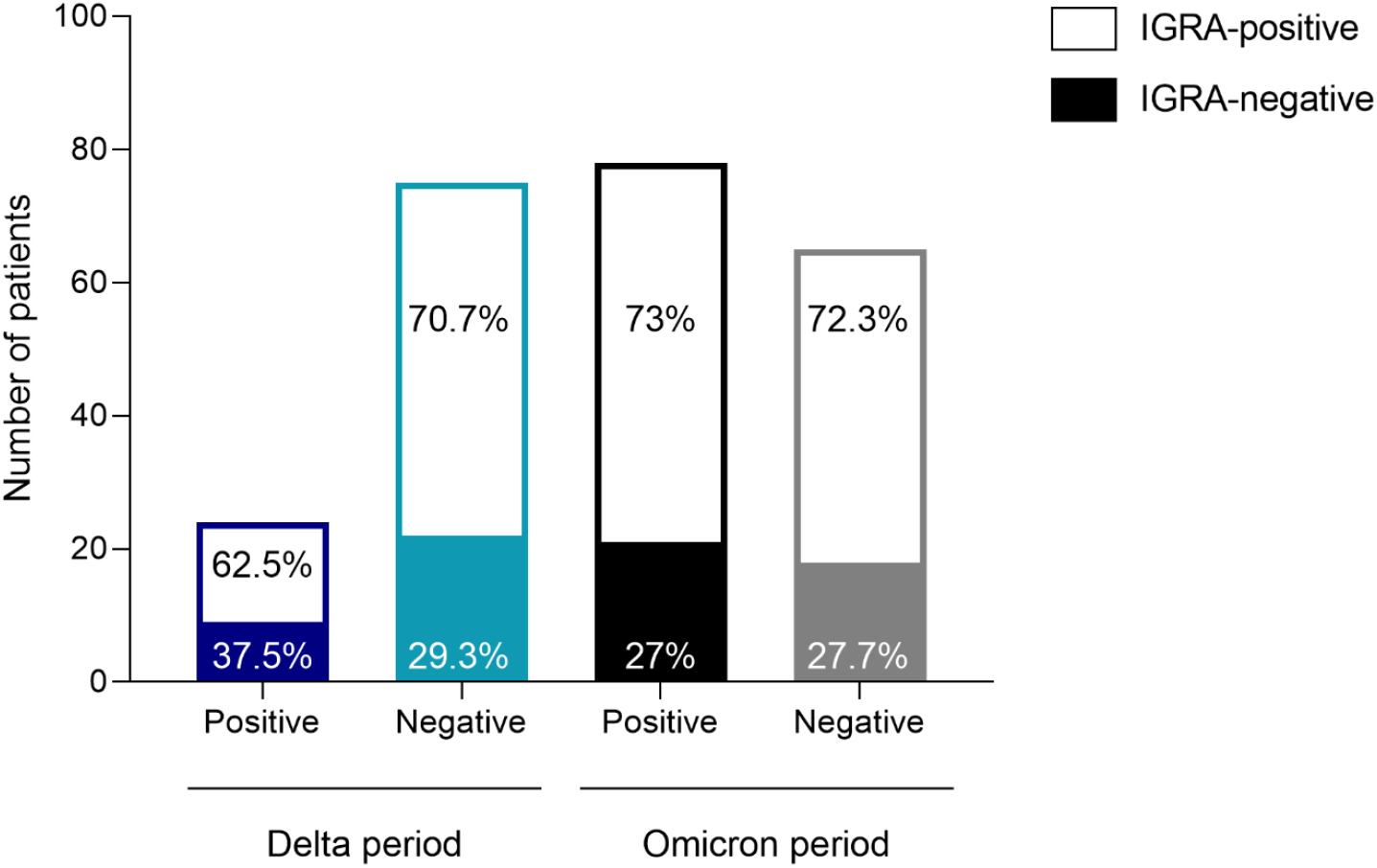
Number of Interferon-gamma release assay (IGRA)-positive or IGRA-negative patients among the positive and negative patients during the Delta and Omicron periods. Numbers in the box represent the percentage of IGRA-positive or negative individuals for each group.

### 3.5. Viral load

No significant difference was observed in terms of normalised viral load between Delta (median = 5.7, IQR [4.4-6.5], log_10_ cp/10^4^ cells) and Omicron patients (median = 5.6, IQR [4.9-6.3], log_10_ cp/10^4^ cells, *p* > 0.05; Suppl. Fig. 4). A moderate negative correlation was observed between normalised viral load and anti-RBD IgG levels during the Delta period (r = -0.3731, *p* = 0.0008, Suppl. Fig. 5), and there was no significant correlation between these two parameters for samples during the Omicron period (r = -0.02737, *p* = 0.7725, Suppl. Fig. 5).

## 4. Discussion

The present study included vaccinated outpatients with symptoms compatible with SARS-CoV-2 infection with the aim to investigate determinants of protection during the periods of Delta and Omicron BA.1 circulation. We found that high circulating levels of anti-RBD IgG and a hybrid infection-vaccination immunisation were associated with a lower risk of symptomatic SARS-CoV-2 infection with the Delta variant. However, this association was not observed in the case of Omicron, except for individuals with hybrid immunity and the highest anti-RBD IgG titres.

We initially compared the clinical characteristics between SARS-CoV-2 positive and negative patients, and between the periods investigated. Among the symptoms recorded in Delta-infected patients, anosmia and ageusia were significantly more frequent than among SARS-CoV-2 negative patients included during the same period. Among Omicron BA.1-infected patients, as in symptomatic individuals not infected with SARS-CoV-2 during the Delta period, more cases reported sore throat than among those infected with Delta variant. These clinical findings are in line with that reported in previous studies (19-21).

Higher titres of anti-spike and anti-RBD IgG in uninfected controls compared to breakthrough cases have proved to decrease the risk of symptomatic COVID-19 mainly with Alpha (1, 4) and Delta variants (5, 7, 22). In the present study, we observed a decrease in the risk of infection as anti-RBD IgG levels increased after adjusting for immunisation regimen in the Delta period. Feng *et al*. previously estimated for the 506 BAU/ml threshold a vaccine efficacy after two doses of ChadOx1nCoV-19 (AZD1222) of 80% against symptomatic infection with Alpha variant (1). We estimated a similar risk of infection during the Delta period for the homologous (two mRNA doses) and heterologous (one dose of a viral vector vaccine and one dose of an mRNA vaccine) vaccination regimens when we applied the same threshold, or even about three-times lower for hybrid immunity group. However, conflicting results have been reported in the case of Omicron variant regarding the value of anti-RBD IgG as a CoP (7-9, 23). The reduction in neutralisation titres observed for Omicron compared to previous variants (24) could indicate that infection can even occur shortly after the booster dose despite high binding antibody levels (25), as observed in the present study. The risk of infection during the Omicron period in the present study was below 20% only in the hybrid group when anti-RBD IgG values were above 5000 BAU/ml, suggesting that the level of anti-spike IgG have a limited impact as predictor of protection (7). In this sense, it is crucial that the anti-spike or anti-RBD IgG protective threshold determined for previous variants is continuously reviewed and adapted to the emerging new variants, which we speculate to be higher or with a lower predictive value as for the previous variants.

The combination of natural immunity and vaccination is attracting more attention and importance due to the globally increasing SARS-CoV-2 infected and vaccinated population. In the present study, we found higher anti-RBD IgG titres in the individuals with hybrid immunity compared to vaccinated patients regardless of the period, as observed elsewhere (26, 27). Regarding the cellular response, a greater proportion of IGRA-positive patients was observed among those with hybrid immunity in the present study, as described previously for the T cell response using IFNγ ELISpot assays (28). These findings are in line with those of several studies that suggested that hybrid immunity can confer more effective cross-variant neutralisation and long-lasting immunity (29-31).

Positive and negative patients with high anti-RBD IgG titres during both periods had high neutralising capacity against the isolate corresponding to their respective period, however, no difference was detected in the neutralising antibody levels between positive and negative patients. These findings might suggest that Delta and Omicron breakthrough infections in patients with high anti-RBD IgG levels are not explained by a lack of neutralising capacity. However, among sera with neutralisation capacity, both samples from Delta and Omicron periods had lower neutralisation capacity against Omicron isolate compared to Delta isolate despite higher anti-RBD IgG levels among those from the Omicron period, which is consistent with that reported by Ai *et al*. (32). This could indicate that neutralising titres against the Omicron variant may not reliably predict at the individual level the risk of SARS-CoV-2 infection (6, 23, 33), conversely to that described for previous VOCs (34). Nevertheless, we could not conclude as to the value of neutralising antibodies as a CoP for Omicron BA.1 variant in the present study as we selected a subset of patients with high anti-RBD IgG levels for the analysis of neutralising antibodies.

In the present study we did not find a significant difference between the proportion of IGRA-positive and -negative patients during Omicron and Delta periods, indicating that this technique was not useful as an immunological marker associated with risk of infection when used qualitatively. However, IGRA-positive patients had higher anti-RBD IgG levels than IGRA-negative patients, which was indirectly associated with a lower risk of infection in the Delta period. In this regard, previous studies have found correlations between IFN-γ release and anti-spike IgG levels in individuals vaccinated with BNT162b2 (35) or in COVID-19 recovered patients (36), although it has not yet been evaluated in the context of hybrid immunity or after the booster dose. These results could suggest that the determination of anti-RBD IgG together with the IFN-γ release could be valuable strategy to predict protection. However, we only assessed the IGRA qualitatively in the present study and, to date, no threshold for the IGRA associated with the risk of infection or the correlation with protection has been estimated.

The viral load in Omicron BA.1 patients was not higher than that in Delta patients, as previously reported in Omicron BA.1 and BA.2 cases (37). In this sense, the higher number of Omicron breakthrough infections with high levels of binding antibodies compared to Delta observed in the present study could not be explained by a higher viral load. There was a weak inverse correlation between viral load and anti-RBD IgG titres among Delta samples while no significant correlation was observed for Omicron samples. The results in Delta-infected patients are only partially in line with that reported by Kim *et al*. who found also an inverse relationship 5-7 days after diagnosis in vaccinated patients with Delta infection (38). However, they did not find differences in cycle threshold (Ct) values according to antibody levels at the time of diagnosis. These findings suggest an association of antibody levels and viral load in breakthrough infections and enhanced viral clearance in patients with higher antibody levels, at least in Delta infections, although it seems to depend on the time of diagnosis after the onset of symptoms.

It is important to note some limitations of the present study. Firstly, many patients were excluded from the analyses to limit heterogeneity of population characteristics, reducing the sample size for some immunisation regimens. In this sense, we grouped the patients into three main immunisation regimens (homologous, heterologous and hybrid), who mainly had two immunisations during the Delta period and three during Omicron period, but some of them had two, three or even four immunisations. It should also be noted that we did not distinguish between the type of mRNA vaccine in the homologous and heterologous regimens (BNT162b2 and mRNA-1273 vaccines). Variability in the number of immunisations may have had an effect on the results and should be evaluated in future studies in a larger number of patients. However, despite these limitations, as we have used a test negative design, the cases and controls had similar characteristics minimizing confounding by health care-seeking behaviour. With regard to neutralisation assays, not all samples were tested for the determination of neutralising capacity since it is a logistically demanding process as it requires live viruses manipulated in a biosafety level-3 laboratory that requires trained staff and specific equipment. Likewise, PRNT_50_ was estimated by the evaluation of samples by eye and microscope and thus is highly dependent on the operator, which may also influence the results.

In conclusion, the present study suggests that higher circulating levels of anti-RBD IgG and hybrid immunity were independently associated with a lower risk of symptomatic SARS-CoV-2 infection in outpatients with differences according to the infecting variant.

## Data Availability

All data produced in the present study are available upon reasonable request to the authors

## Acknowledgements

We thank all the staff members of the occupational health and medicine department of the *Hospices Civils de Lyon* who contributed to the sample collection. We thank all the technicians from the virology laboratory whose work made it possible to obtain all these data. We thank Philip Robinson (DRCI, Hospices Civils de Lyon) for his help in manuscript preparation. Lastly, we thank all the healthcare workers for their participation in this clinical study.

## Author contributions

A.R., C.S., L.J., B.C., F.M., R.H., B.P., B.L., M.-A.T., S.T.-A. and A.B. conceived and design the study. A.R., C.S., L.J., M.A.T., S.T.-A. and A.B. analysed and interpreted the data. G.D., M.V., V.I., A.O., M.D., C.G., K.B.-P., L.G. and K.S. conducted and managed experimental work. A.R. and C.S. wrote the first draft of the manuscript. All authors critically reviewed and approved the final version of the manuscript.

## Ethics statement

The study is registered on ClinicalTrials.gov (NCT05060939). Written informed consent was obtained from all participants and approval was obtained from the regional review board in July 2021 (*Comité de Protection des Personnes Sud Méditerranée I*, Marseille, France; ID-RCB 2021-A01877-34).

---

**Suppl. Table 1.**
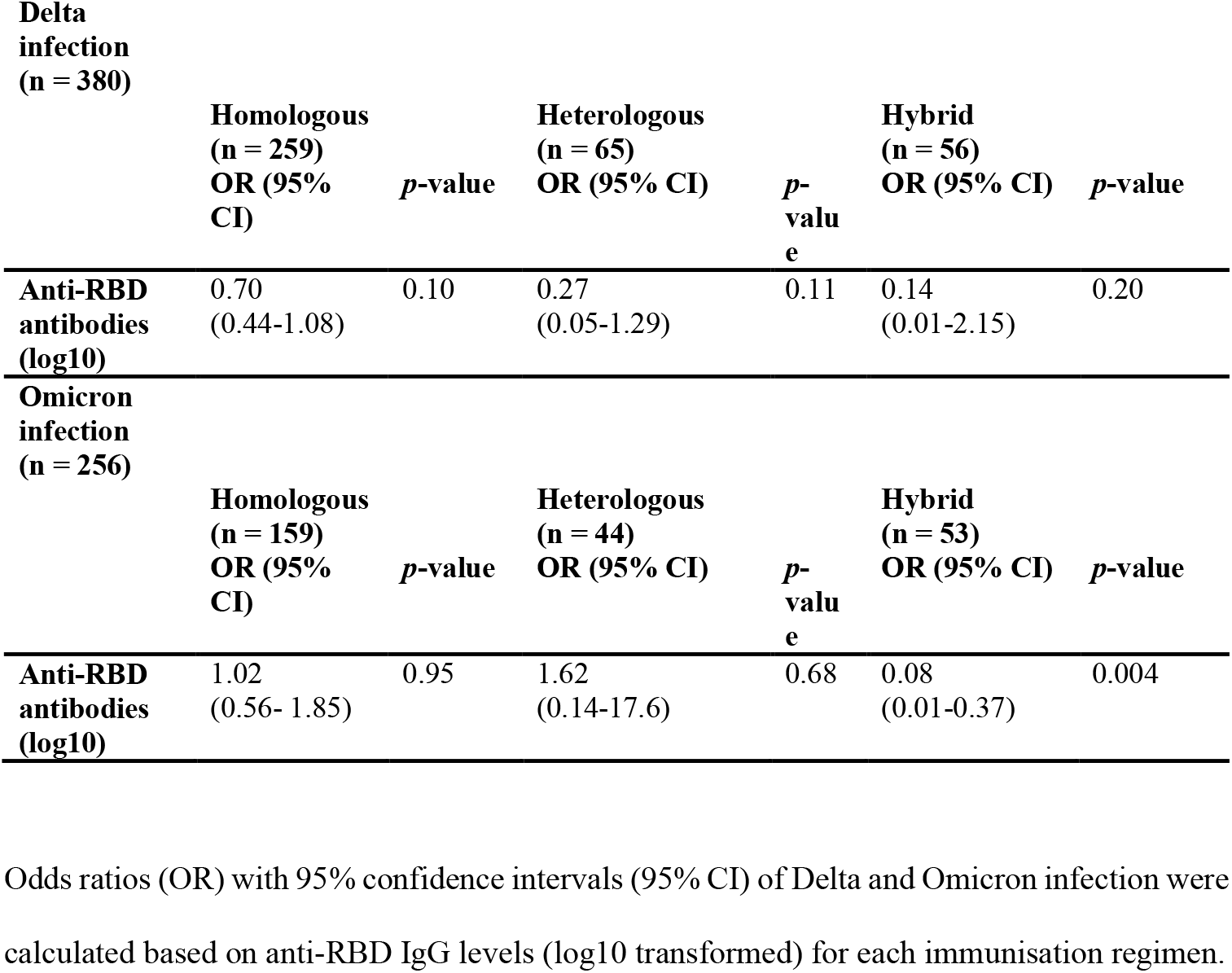
Risk of infection according to anti-RBD IgG levels and stratified by immunization regimen.

**Suppl. Table 2.**
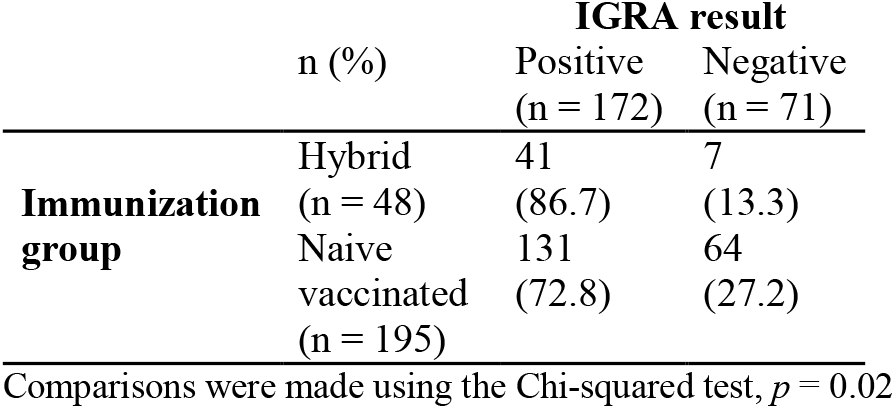
Number and proportion of patients with and without hybrid immunity according to the Interferon-gamma release assay (IGRA) result considering the Delta and Omicron periods together.

**Suppl. Fig. 1.**
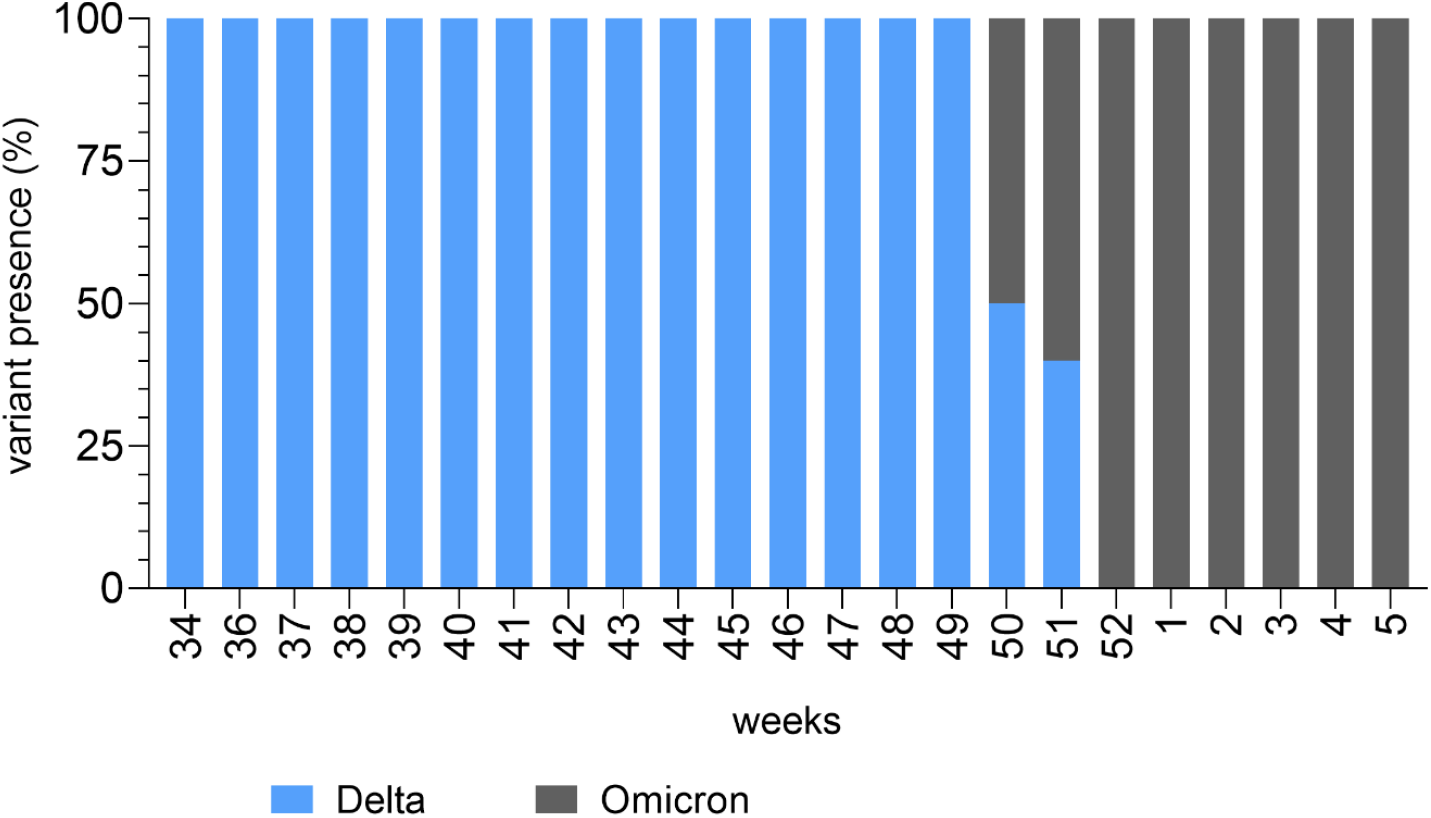
Percentages of detection of each variant among all SARS-CoV-2 positive patients each week from 20 August 2021 (week 34) to 2 February 2022 (week 5). Weeks 34 to 49 corresponded to the Delta period and weeks 52 to 5 corresponded to the Omicron period. Weeks 50-51 were excluded from the analysis as they corresponded to the co-circulation of Delta and Omicron.

**Suppl. Fig. 2.**
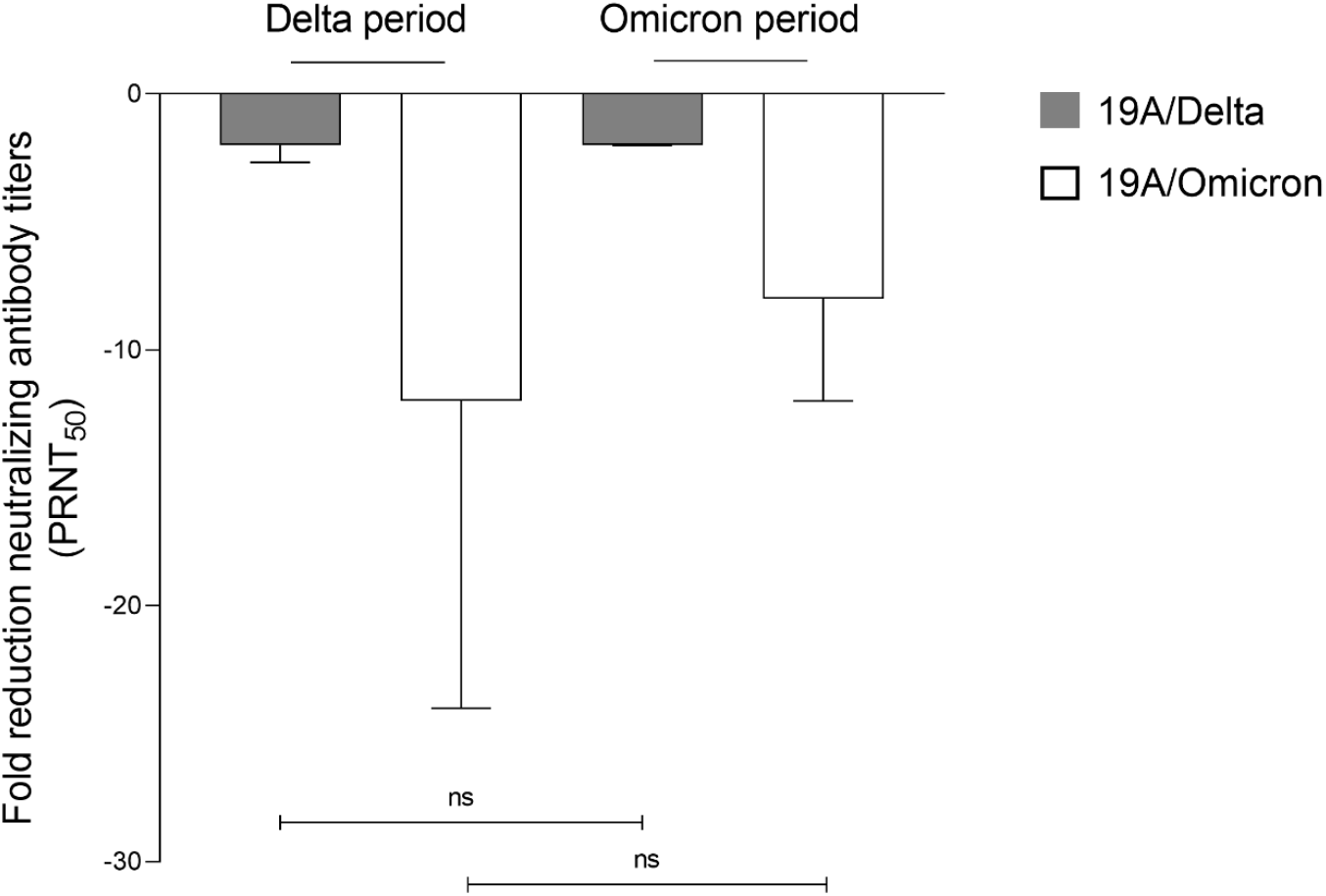
Comparison of neutralising antibody titres of sera collected during the Delta and Omicron periods. A 50% plaque reduction neutralization test (PRNT_50_) was conducted to determine the neutralisation titre of 35 patient samples from both the Delta and Omicron periods against the live 19A, Delta and Omicron isolates. Fold-reduction of neutralisation titres against Delta and Omicron isolate compared to 19A isolate for the Delta and Omicron periods was calculated. Bars represent the median and error bars the 95% confidence interval (CI). Comparisons were made using the Mann-Whitney test. ns, non-significant.

**Suppl. Fig. 3.**
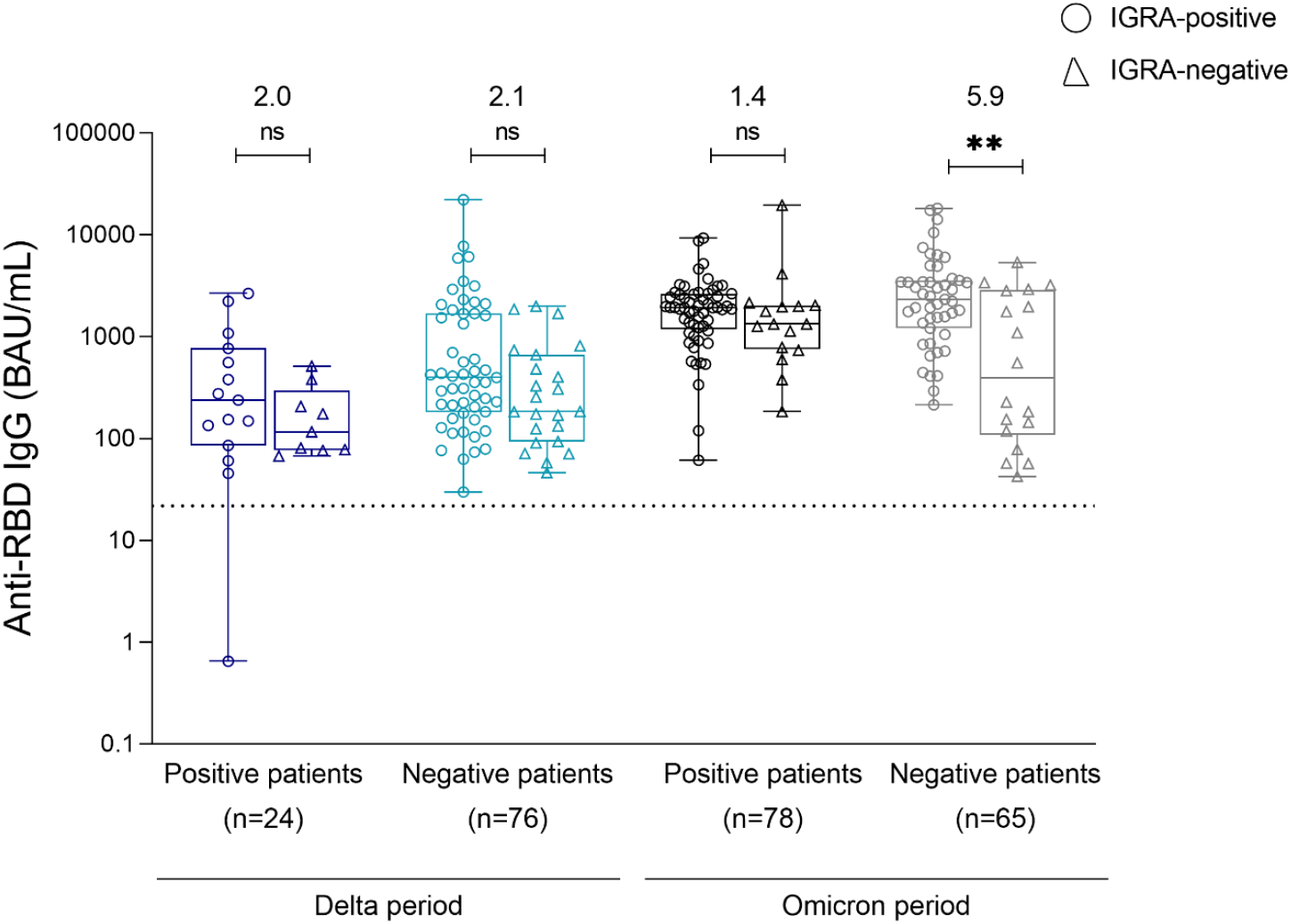
Comparison of anti-RBD IgG levels between Interferon Gamma release assay (IGRA) -positive and IGRA-negative samples for the Delta and Omicron periods. An Interferon Gamma Release Assay (IGRA) was performed to assess T cell function among patients following stimulation with a restricted pool of peptides (RPP) derived from SARS-CoV-2 structural proteins. A chemiluminescence immunoassay (CLIA) was performed in order to determine anti-RBD IgG levels, expressed in binding antibody units (BAU)/mL (n = 243) for IGRA-positive and IGRA-negative samples for the Delta and Omicron periods. The dotted line represents the threshold of positivity (≥ 21.8 BAU/mL). Box plots represent the median and interquartile range [IQR], and whiskers represent the range. Circles represent IGRA-positive samples and triangles represent IGRA-negative samples. Comparisons were made using the Kruskal-Wallis test; ns: non-significant, \*\**p*<0.01. Fold differences between IGRA-positive and IGRA-negative samples are indicated above the graph.

**Suppl. Fig. 4.**
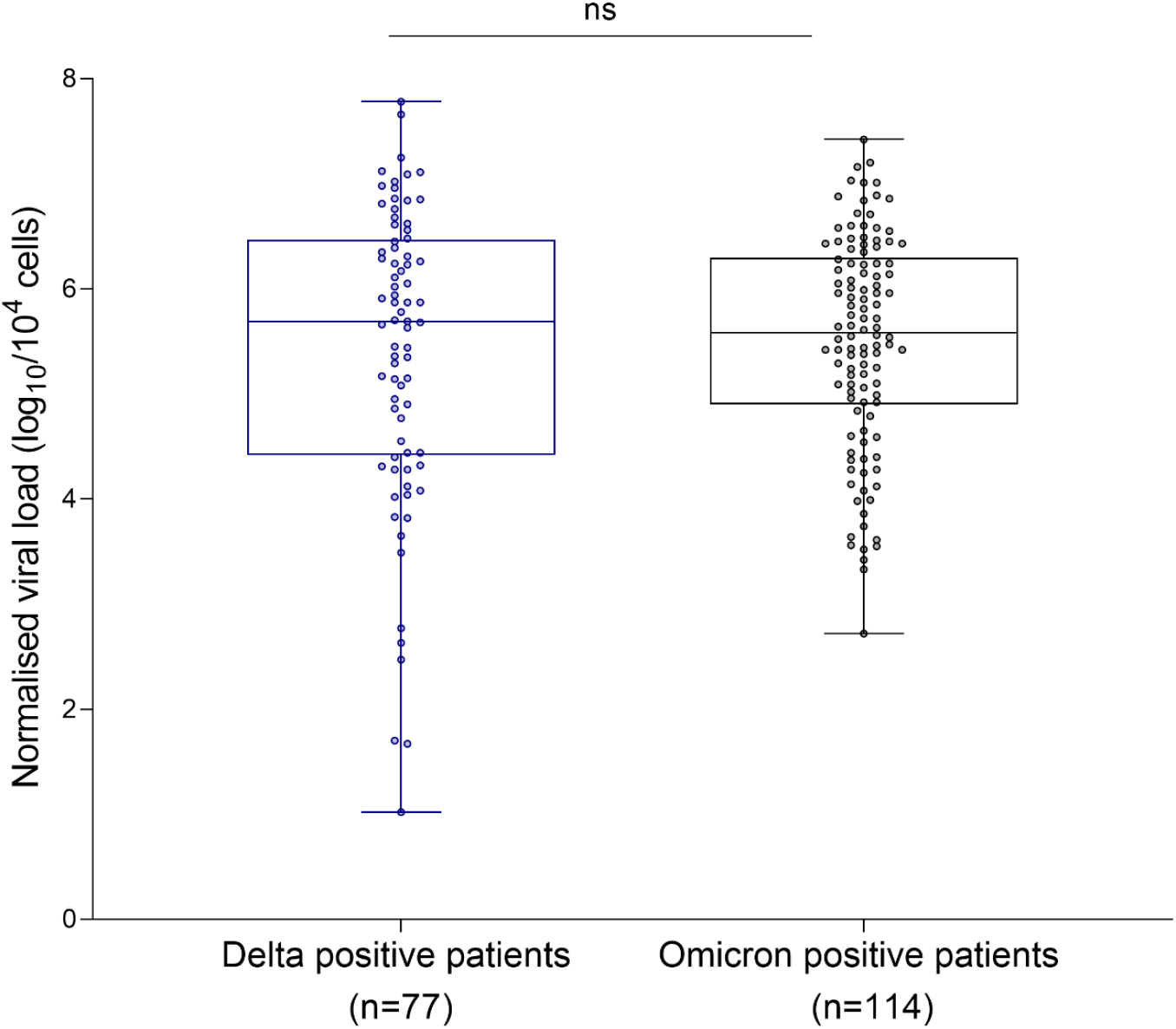
Normalised viral load (log_10_/10^4^ cells) in patients from Delta and Omicron periods. Comparisons were made using the Mann-Whitney test, ns: non-significant.

**Suppl. Fig. 5.**
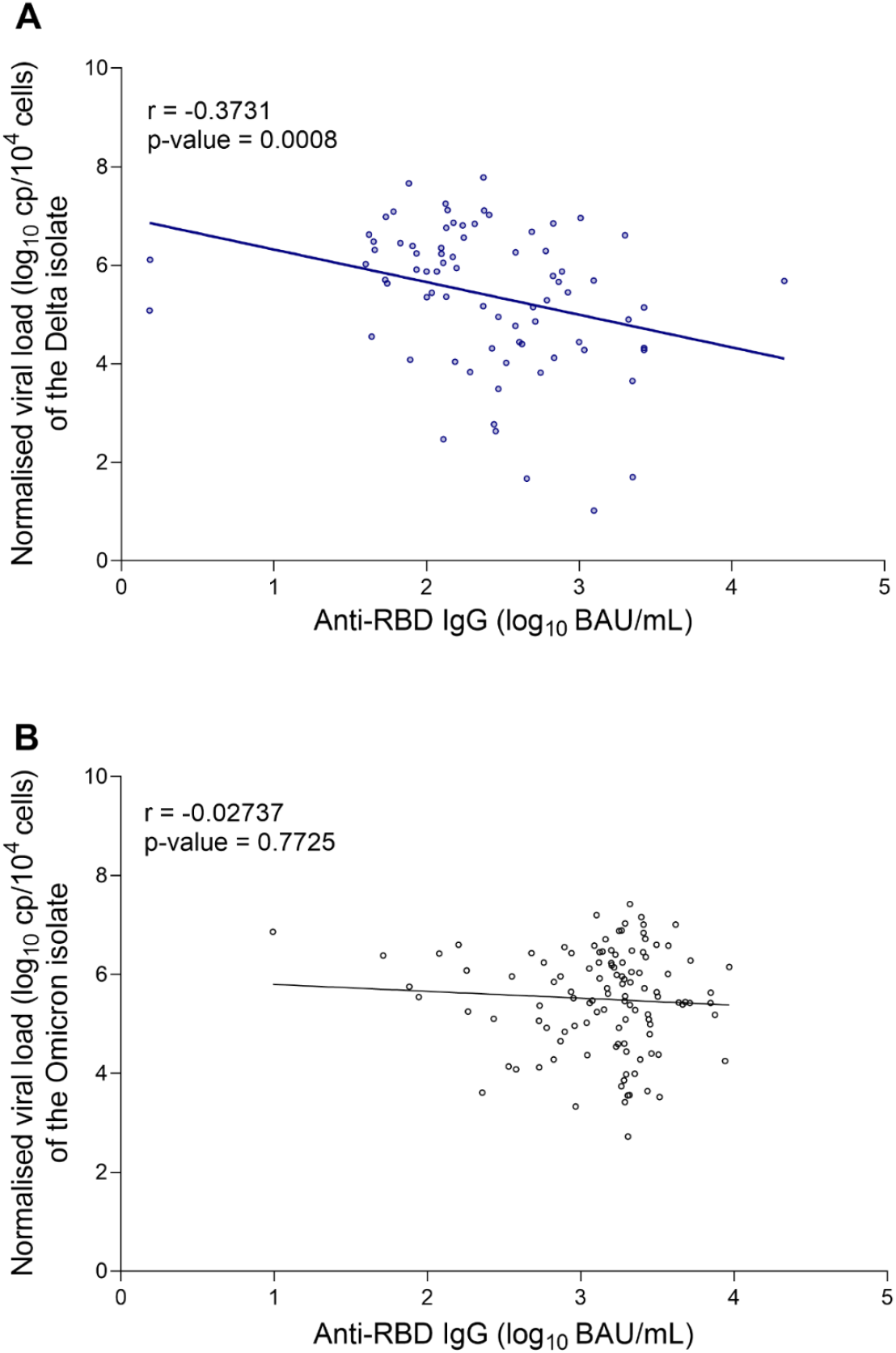
Correlation between anti-RBD IgG levels and normalised viral load. Correlation analysis between anti-RBD IgG levels and normalised viral load for samples corresponding to Delta positive patients (A) and Omicron positive patients (B). Spearman’s rank correlation test was used.

## References

1. Feng S, Phillips DJ, White T, Sayal H, Aley PK, Bibi S, et al. Correlates of protection against symptomatic and asymptomatic SARS-CoV-2 infection. Nat Med. 2021; 27: 2032–40. https://doi.org/10.1038/s41591-021-01540-1.

2. Goldblatt D, Alter G, Crotty S, Plotkin SA. Correlates of protection against SARS-CoV-2 infection and COVID-19 disease. Immunol Rev. 2022. https://doi.org/10.1111/imr.13091.

3. Khoury DS, Cromer D, Reynaldi A, Schlub TE, Wheatley AK, Juno JA, et al. Neutralizing antibody levels are highly predictive of immune protection from symptomatic SARS-CoV-2 infection. Nat Med. 2021; 27: 1205–11. https://doi.org/10.1038/s41591-021-01377-8.

4. Bergwerk M, Gonen T, Lustig Y, Amit S, Lipsitch M, Cohen C, et al. Covid-19 Breakthrough Infections in Vaccinated Health Care Workers. N Engl J Med. 2021; 385: 1474–84. https://doi.org/10.1056/NEJMoa2109072.

5. Wei J, Pouwels KB, Stoesser N, Matthews PC, Diamond I, Studley R, et al. Antibody responses and correlates of protection in the general population after two doses of the ChAdOx1 or BNT162b2 vaccines. Nat Med. 2022; 28: 1072–82. https://doi.org/10.1038/s41591-022-01721-6.

6. Takheaw N, Liwsrisakun C, Chaiwong W, Laopajon W, Pata S, Inchai J, et al. Correlation Analysis of Anti-SARS-CoV-2 RBD IgG and Neutralizing Antibody against SARS-CoV-2 Omicron Variants after Vaccination. Diagnostics (Basel). 2022; 12. https://doi.org/10.3390/diagnostics12061315.

7. Stærke NB, Reekie J, Nielsen H, Benfield T, Wiese L, Knudsen LS, et al. Levels of SARS-CoV-2 antibodies among fully vaccinated individuals with Delta or Omicron variant breakthrough infections. Nat Commun. 2022; 13: 4466. https://doi.org/10.1038/s41467-022-32254-8.

8. Mollura M, Sarti R, Levi R, Pozzi C, Azzolini E, Politi LS, et al. Antibody Titer Correlates with Omicron Infection in Vaccinated Healthcare Workers. Viruses. 2022; 14: 2605. https://doi.org/10.3390/v14122605.

9. Favresse J, Dogné JM, Douxfils J. Assessment of the humoral response in Omicron breakthrough cases in healthcare workers who received the BNT162b2 booster. Clin Chem Lab Med. 2022; 60: e153–e6. https://doi.org/10.1515/cclm-2022-0323.

10. Kent SJ, Khoury DS, Reynaldi A, Juno JA, Wheatley AK, Stadler E, et al. Disentangling the relative importance of T cell responses in COVID-19: leading actors or supporting cast? Nat Rev Immunol. 2022; 22: 387–97. https://doi.org/10.1038/s41577-022-00716-1.

11. Primorac D, Brlek P, Matišić V, Molnar V, Vrdoljak K, Zadro R, et al. Cellular Immunity-The Key to Long-Term Protection in Individuals Recovered from SARS-CoV-2 and after Vaccination. Vaccines (Basel). 2022; 10: 442. https://doi.org/10.3390/vaccines10030442.

12. Tan AT, Lim JM, Le Bert N, Kunasegaran K, Chia A, Qui MD, et al. Rapid measurement of SARS-CoV-2 spike T cells in whole blood from vaccinated and naturally infected individuals. J Clin Invest. 2021; 131. https://doi.org/10.1172/JCI152379.

13. Bal A, Simon B, Destras G, Chalvignac R, Semanas Q, Oblette A, et al. Detection and prevalence of SARS-CoV-2 co-infections during the Omicron variant circulation in France. Nat Commun. 2022; 13: 6316. https://doi.org/10.1038/s41467-022-33910-9.

14. Siemens Healthcare Diagnostics. Atellica IM SARS-CoV-2 IgG (sCOVG) - Instructions for Use 2020.

15. WHO First WHO International Standard for anti-SARS-CoV-2 immunoglobulin. 2020. https://www.nibsc.org/documents/ifu/20-136.pdf. Last accessed on 16 December 2022.

16. Legros V, Denolly S, Vogrig M, Boson B, Siret E, Rigaill J, et al. A longitudinal study of SARS-CoV-2-infected patients reveals a high correlation between neutralizing antibodies and COVID-19 severity. Cell Mol Immunol. 2021; 18: 318–27. https://doi.org/10.1038/s41423-020-00588-2.

17. Bal A, Pozzetto B, Trabaud MA, Escuret V, Rabilloud M, Langlois-Jacques C, et al. Evaluation of High-Throughput SARS-CoV-2 Serological Assays in a Longitudinal Cohort of Patients with Mild COVID-19: Clinical Sensitivity, Specificity, and Association with Virus Neutralization Test. Clin Chem. 2021; 67: 742–52. https://doi.org/10.1093/clinchem/hvaa336.

18. Saade C, Brengel-Pesce K, Gaymard A, Trabaud MA, Destras G, Oriol G, et al. Dynamics of viral shedding during ancestral or Omicron BA.1 SARS-CoV-2 infection and enhancement of pre-existing immunity during breakthrough infections. Emerg Microbes Infect. 2022; 11: 2423–32. https://doi.org/10.1080/22221751.2022.2122578.

19. Pacchiarini N, Sawyer C, Williams C, Sutton D, Roberts C, Simkin F, et al. Epidemiological analysis of the first 1000 cases of SARS-CoV-2 lineage BA.1 (B.1.1.529, Omicron) compared with co-circulating Delta in Wales, UK. Influenza Other Respir Viruses. 2022; 16: 986–993. https://doi.org/10.1111/irv.13021.

20. Menni C, Valdes AM, Polidori L, Antonelli M, Penamakuri S, Nogal A, et al. Symptom prevalence, duration, and risk of hospital admission in individuals infected with SARS-CoV-2 during periods of omicron and delta variant dominance: a prospective observational study from the ZOE COVID Study. Lancet. 2022; 399: 1618–24. https://doi.org/10.1016/S0140-6736(22)00327-0.

21. Lippi G, Nocini R, Henry BM. Analysis of online search trends suggests that SARS-CoV-2 Omicron (B.1.1.529) variant causes different symptoms. J Infect. 2022; 84: e76–e7. https://doi.org/10.1016/j.jinf.2022.02.011.

22. Aldridge RW, Yavlinsky A, Nguyen V, Eyre MT, Shrotri M, Navaratnam AMD, et al. SARS-CoV-2 antibodies and breakthrough infections in the Virus Watch cohort. Nat Commun. 2022; 13: 4869. https://doi.org/10.1038/s41467-022-32265-5.

23. Lee J, Park S, Kim JY, Lim SY, Chang E, Bae S, et al. No correlation of neutralizing antibody titers against the Omicron variant after a booster dose of COVID-19 vaccines with subsequent breakthrough Omicron infections among healthcare workers. J Infect. 2022; 85: e177–e80. https://doi.org/10.1016/j.jinf.2022.10.007.

24. Planas D, Saunders N, Maes P, Guivel-Benhassine F, Planchais C, Buchrieser J, et al. Considerable escape of SARS-CoV-2 Omicron to antibody neutralization. Nature. 2022; 602: 671–5. https://doi.org/10.1038/s41586-021-04389-z.

25. Cui Z, Liu P, Wang N, Wang L, Fan K, Zhu Q, et al. Structural and functional characterizations of infectivity and immune evasion of SARS-CoV-2 Omicron. Cell. 2022; 185: 860-71.e13. https://doi.org/10.1016/j.cell.2022.01.019.

26. Zar HJ, MacGinty R, Workman L, Botha M, Johnson M, Hunt A, et al. Natural and hybrid immunity following four COVID-19 waves: A prospective cohort study of mothers in South Africa. EClinicalMedicine. 2022; 53: 101655. https://doi.org/10.1016/j.eclinm.2022.101655.

27. Epsi NJ, Richard SA, Lindholm DA, Mende K, Ganesan A, Huprikar N, et al. Understanding ‘hybrid immunity’: comparison and predictors of humoral immune responses to SARS-CoV-2 infection and COVID-19 vaccines. Clin Infect Dis. 2022. https://doi.org/10.1093/cid/ciac392.

28. Naranbhai V, Nathan A, Kaseke C, Berrios C, Khatri A, Choi S, et al. T cell reactivity to the SARS-CoV-2 Omicron variant is preserved in most but not all individuals. Cell. 2022; 185: 1041-51.e6. https://doi.org/10.1016/j.cell.2022.01.029.

29. Suryawanshi R, Ott M. SARS-CoV-2 hybrid immunity: silver bullet or silver lining? at Rev Immunol. 2022: 1–2. https://doi.org/10.1038/s41577-022-00771-8.

30. Wang Z, Muecksch F, Schaefer-Babajew D, Finkin S, Viant C, Gaebler C, et al. Naturally enhanced neutralizing breadth against SARS-CoV-2 one year after infection. Nature. 2021; 595: 426–31. https://doi.org/10.1038/s41586-021-03696-9.

31. Hall V, Foulkes S, Insalata F, Kirwan P, Saei A, Atti A, et al. Protection against SARS-CoV-2 after Covid-19 Vaccination and Previous Infection. N Engl J Med. 2022; 386: 1207–20. https://doi.org/10.1056/NEJMoa2118691.

32. Ai J, Zhang H, Zhang Y, Lin K, Zhang Y, Wu J, et al. Omicron variant showed lower neutralizing sensitivity than other SARS-CoV-2 variants to immune sera elicited by vaccines after boost. Emerg Microbes Infect. 2022; 11: 337–43. https://doi.org/10.1080/22221751.2021.2022440.

33. Torres I, Giménez E, Albert E, Zulaica J, Álvarez-Rodríguez B, Burgos JS, et al. SARS-CoV-2 Omicron BA.1 variant breakthrough infections in nursing home residents after an homologous third dose of the Comirnaty® COVID-19 vaccine: Looking for correlates of protection. J Med Virol. 2022; 94: 4216–23. https://doi.org/10.1002/jmv.27867.

34. Cromer D, Steain M, Reynaldi A, Schlub TE, Wheatley AK, Juno JA, et al. Neutralising antibody titres as predictors of protection against SARS-CoV-2 variants and the impact of boosting: a meta-analysis. The Lancet Microbe. 2022; 3: e52–e61. https://doi.org/10.1016/S2666-5247(21)00267-6.

35. Kurteva E, Vasilev G, Tumangelova-Yuzeir K, Ivanova I, Ivanova-Todorova E, Velikova T, et al. Interferon-gamma release assays outcomes in healthy subjects following BNT162b2 mRNA COVID-19 vaccination. Rheumatol Int. 2022; 42: 449–56. https://doi.org/10.1007/s00296-022-05091-7.

36. Schiffner J, Backhaus I, Rimmele J, Schulz S, Möhlenkamp T, Klemens JM, et al. Long-Term Course of Humoral and Cellular Immune Responses in Outpatients After SARS-CoV-2 Infection. Front Public Health. 2021; 9: 732787. https://doi.org/10.3389/fpubh.2021.732787.

37. Yuasa S, Nakajima J, Takatsuki Y, Takahashi Y, Tani-Sassa C, Iwasaki Y, et al. Viral load of SARS-CoV-2 Omicron is not high despite its high infectivity. J Med Virol. 2022; 94: 5543–6. https://doi.org/10.1002/jmv.27974.

38. Kim MH, Nam Y, Son NH, Heo N, Kim B, Kang E, et al. Antibody Level Predicts the Clinical Course of Breakthrough Infection of COVID-19 Caused by Delta and Omicron Variants: A Prospective Cross-Sectional Study. Open Forum Infect Dis. 2022;9: ofac262. https://doi.org/10.1093/ofid/ofac262.

